# Standardized country-level delivery unit cost estimates for routine childhood, routine adolescent, and campaign vaccination: an updated modeling analysis

**DOI:** 10.64898/2026.03.15.26348434

**Authors:** Allison Portnoy, Emma Clarke-Deelder, Taylor A. Holroyd, Daniel R. Hogan, Alex Olateju Adjagba, Logan Brenzel, Bryan N. Patenaude, So Yoon Sim, Tewodaj Mengistu, Nicolas A. Menzies

## Abstract

**Background:** Reliable estimates of immunization delivery costs are essential for planning, budgeting, and economic evaluation. Despite recent empirical studies, many low- and middle-income countries (LMICs) lack up-to-date cost estimates, particularly for adolescent and campaign vaccination. This study generated standardized country-level delivery cost estimates across vaccination modalities in LMICs.

**Methods:** Using data from empirical costing studies in the 2024 update of the Immunization Delivery Cost Catalogue, we fitted Bayesian meta-regression models to estimate per-dose delivery costs, not including vaccine and injection supplies costs, for routine childhood vaccination, routine adolescent vaccination (using human papillomavirus vaccination as a proxy), and mass campaigns. Models incorporated country-level covariates (gross domestic product per capita, population size, diphtheria-tetanus-pertussis third-dose coverage, urbanization, population density, and under-five mortality) and study-level characteristics (cost category and costing perspective). Fitted models were used to generate country-specific and population-weighted mean economic and financial costs per dose in 2024 US dollars.

**Results:** The analysis included 141 observations for routine childhood, 49 for adolescent, and 104 for campaign vaccination. The population-weighted mean full economic cost per dose across LMICs was $4.27 (95% prediction interval: $1.95–11.58) for routine childhood vaccination, $20.56 ($8.56–57.83) for adolescent vaccination, and $14.71 ($4.90–86.98) for campaigns. Corresponding estimates for incremental economic costs per dose were $16.64 ($7.90–38.22), $20.43 ($8.27–47.78), and $3.53 ($1.70–8.56). Substantial heterogeneity was observed across countries, delivery modalities, and cost perspectives.

**Conclusions:** Leveraging an expanded evidence-base and Bayesian meta-regression, this study provides updated delivery cost estimates across vaccination modalities for 136 LMICs. These estimates offer a practical resource for analyses where empirical cost data are unavailable.

**Key points:**

1. This study provides updated estimates of how much it costs to deliver vaccines across low- and middle-income countries, showing that costs vary widely depending on how vaccines are delivered and the context of each country.
2. On average, delivering vaccines to adolescents (facility-based, school-based, and outreach) is substantially more expensive than routine childhood vaccination, while campaign delivery costs fall in between, reflecting differences in logistics and delivery strategies.
3. These results offer practical, comparable cost benchmarks that policymakers and researchers can use to plan immunization programs and make informed decisions when country-specific data are not available.

## 1. Background

Routine immunization remains a cornerstone of primary health care, and impacts 14 of the 17 Sustainable Development Goals (SDGs) adopted in 2015 [1, 2]. In the decade since, the global immunization landscape has seen increasing access to newer, higher-cost vaccines—especially pneumococcal conjugate vaccine (PCV) and rotavirus vaccine—and renewed efforts to scale up human papillomavirus (HPV) vaccination [3–6]. In addition to routine services, preventive vaccination campaigns are periodically implemented to close immunity gaps (e.g., measles-containing vaccine follow-up campaigns) or rapidly increase population-level immunity (e.g., measles–rubella catch-up campaigns). Despite this, indicators of vaccine coverage have either decreased or plateaued in recent years. For example, global third dose coverage of diphtheria-tetanus pertussis vaccine (DTP3) has varied around 85%, and measles coverage has not fully returned to pre-pandemic levels [7–9].

The Immunization Agenda 2030 (IA2030), launched in 2021, reaffirmed immunization as a key driver of health equity, pandemic preparedness, and sustainable development, while emphasizing a life-course approach to vaccination that includes adolescence and older age groups [5]. However, implementing IA2030 requires robust financial planning and reliable evidence on vaccine delivery costs. The need for this evidence is amplified by growing fiscal constraints on global health programs in the mid-2020s, including reductions in external development assistance and constrained domestic health budgets [10]. These reductions in official development assistance for health from major donor countries have reduced the predictability and volume of external financing that has historically subsidized immunization delivery in many low-income settings [11–13]. Concurrently, several countries are progressing through Gavi’s co-financing transition process, which requires governments to assume an increasing share of immunization program costs as Gavi support is phased out, while some countries face constraints on their ability to mobilize sufficient additional domestic health financing [14]. These dynamics have been documented in recent analyses of country-level development assistance and fiscal space [4, 10, 15].

In this context, the availability of current, modality-specific, country-level delivery cost estimates is particularly valuable. Countries that can demonstrate the full financial cost of sustaining immunization coverage are better positioned to justify budget protection within domestic resource allocation processes. Similarly, donors and multilateral agencies providing transitional financing, i.e., time-limited external support as countries assume increasing responsibility for financing their immunization programs, require reliable cost benchmarks to assess the realism of government financing commitments and design appropriate support mechanisms. Such estimates can inform decisions about financial sustainability, enable efficiency analyses, and support applications for external assistance from mechanisms such as Gavi, the Vaccine Alliance.

Despite notable progress—including the expansion of multi-country databases such as the Immunization Delivery Cost Catalogue (IDCC) [16]—evidence gaps persist. Many LMICs still rely on outdated or non-representative cost data, limiting their ability to plan, budget, and evaluate immunization programs effectively. Continued investment in empirical costing studies remains critical to inform policy decisions and sustain immunization impact, but the challenging fiscal environment and budget constraints likewise impact the feasibility of conducting these studies [4]. Until locally-generated costing evidence becomes available for all countries, estimates extrapolated from other settings remain valuable for filling evidence gaps, demonstrated by the multiple uses of earlier estimates on which the current study builds [17–19].

The objective of this study was to update previously published estimates of standardized country-level delivery costs of vaccination for all LMICs [20] in three ways: (1) incorporate newly-available data [16]; (2) estimate outcomes for routine adolescent and campaign vaccine delivery; and (3) revise the estimation framework with more robust measures of model fit. We present estimates of delivery cost per dose across LMICs, by World Bank income level, by WHO region, and by country.

## 2. Methods

Using the available study-level empirical data on per-dose delivery costs of vaccination, we developed a Bayesian meta-regression model to predict unit costs for routine fixed facility and outreach delivery of childhood vaccines (i.e., under-five), routine fixed facility and outreach delivery of adolescent vaccines (using HPV vaccine as a proxy), and vaccine delivery via mass campaigns (excluding vaccines and supplies such as syringes and safety boxes) in LMICs, according to World Bank country income classification in 2026 [21], updating a prior analysis [20]. In this section, we describe the data used to fit the model and the revised modeling methods for the updated analysis.

### 2.1 Study data

We relied on a publicly available database describing immunization delivery costs in LMIC settings—the Immunization Delivery Cost Catalogue (IDCC) maintained by the Immunization Costing Action Network (ICAN) [22, 23]. The IDCC is an online web catalog and downloadable Excel spreadsheet of immunization delivery cost evidence in LMIC settings, which describes the results of a systematic review of published and grey literature (covering three categories of key words: “immunization” AND “cost” AND “delivery”) available between January 2005 and December 2023. The search of the peer-reviewed literature included six major electronic databases, including EconLit, Embase, Medline (via PubMed), NHS-EED, Web of Science, and WHO Global Index Medicus. Search terms included three categories of keywords— “immunization” AND “cost” AND delivery”—translated into the query language of each database. All resources with full text availability in English, French, or Spanish, conducted in LMIC settings, that included a form of delivery unit cost data from primary data collection were included. From the IDCC, we identified studies that reported delivery costs of routine (i.e., fixed facility and outreach) childhood vaccination (i.e., vaccines delivered up to age five), routine adolescent vaccination (using HPV vaccination as a proxy, including ages 9–15 and both school-based and facility-based delivery), and campaign vaccination. Delivery costs included labor, supply chain, capital, and other service delivery costs, and excluded costs related to the vaccine product, injection supplies, and wastage. “Labor” includes the costs of personnel administering vaccinations (as well as the value of volunteer labor for economic costs); “supply chain” includes costs for cold chain equipment, vehicles, transport, and fuel; “capital” includes either the annualized value of capital investments when estimating economic costs or expenditures on capital equipment when estimating financial costs; and “other service delivery” includes costs for program management (i.e., supervision and monitoring), training, social mobilization, and disease surveillance (Appendix Table A; appendices can be found in the Electronic Supplementary Material) [24]. We excluded studies that did not report a cost per dose, studies for which a cost per dose could not be calculated, and studies that did not define the cost categories included in the cost per dose. We also excluded studies that focused targeting vaccine delivery by factors other than age group or school year (i.e., high-risk populations) and studies that could not be categorized into either economic or financial costs.

For the identified studies, we extracted the study year, estimates of the delivery cost per dose, whether the vaccine was delivered through a campaign or routine delivery modality [*Routine*], whether the costing was for the full vaccination or incremental costs of adding a new antigen [*Full*], and whether the presented costs were from the financial or economic perspective [*Econ*]. For routine delivery (both childhood and adolescent vaccines), full costing represents the ongoing (i.e., recurrent) costs associated with vaccine delivery whereas incremental costing represents both the start-up (i.e., introduction) and recurrent delivery costs associated with adding a vaccine to the national immunization program. For campaign delivery, full costing represents the total operational and system costs required to conduct a campaign (i.e., an average cost per dose) whereas incremental costing represents the additional resources required to execute the campaign, conditional on existing system capacity (i.e., marginal cost per dose). Financial costs refer to accounting transactions (i.e., expenditures or monetary outlays) whereas economic costs include both financial costs as well as the value of resources based on their opportunity cost, regardless of whether a financial transaction occurred (e.g., community outreach workers’ volunteer labor) [25]. If any information reported in the IDCC was unclear for this analysis, we confirmed the information in the original studies. For the observed cost per dose outcome, we extracted the observation with the highest level of granularity available from a given study. For example, if the total vaccine delivery cost per dose were also reported as individual cost categories (i.e., labor, supply chain, capital, and other service delivery), we extracted the cost data for each component.

The IDCC 2024 update included delivery cost per dose estimates in 2022 US dollars. In order to present estimates in 2024 US dollars, all cost estimates were first expressed in local currency units (LCU), inflated from the study year to 2024 using the country-specific consumer price index (CPI) [26], and then converted to 2024 US dollars using the 2024 LCU-to-USD exchange rate [27]. Estimates originally reported in US dollars were first converted to LCU using the study-year exchange rate before applying the local CPI adjustment and subsequent conversion to 2024 US dollars.

Based on previous work, we compiled data on covariates potentially associated [17, 20] with immunization delivery unit costs and with estimates available for all 136 LMICs: the year of data collection (*Year*), log gross domestic product (GDP) per capita (log(*GDP*)); reported diphtheria-tetanus-pertussis third dose coverage (*DTP3*); log under-five population (log(*Pop*)); log under-five mortality rate (log(*U5MR*)); log population density (log(*Density*)); and percentage urban population (*Urban*) [28, 29]. Model covariates were compiled for the year of data collection for each study. By using a log transformation of specified covariates, we assumed these explanatory factors relate to the outcome on the multiplicative scale rather than linearly (for example, a doubling in per capita GDP produces a fixed increase in the outcome).

### 2.2 Prediction model

We used a Bayesian meta-regression model to regress immunization delivery unit costs against country-level and study-level explanatory variables separately for full costing studies and incremental costing studies (Table 1). Continuous variables (*Year*, log(*GDP*), *DTP3*, log(*Pop*), log(*U5MR*), log(*Density*), *Urban*) were standardized to mean zero and unit standard deviation before fitting the regression model. We adopted an analytic model that allowed the synthesis of cost estimates that included different combinations of cost categories. Under this approach, a regression equation with a separate cost intercept was specified for each of four cost categories, i.e., labor 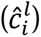, supply chain 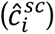, service delivery 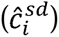, and capital 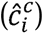:

**Table 1.**
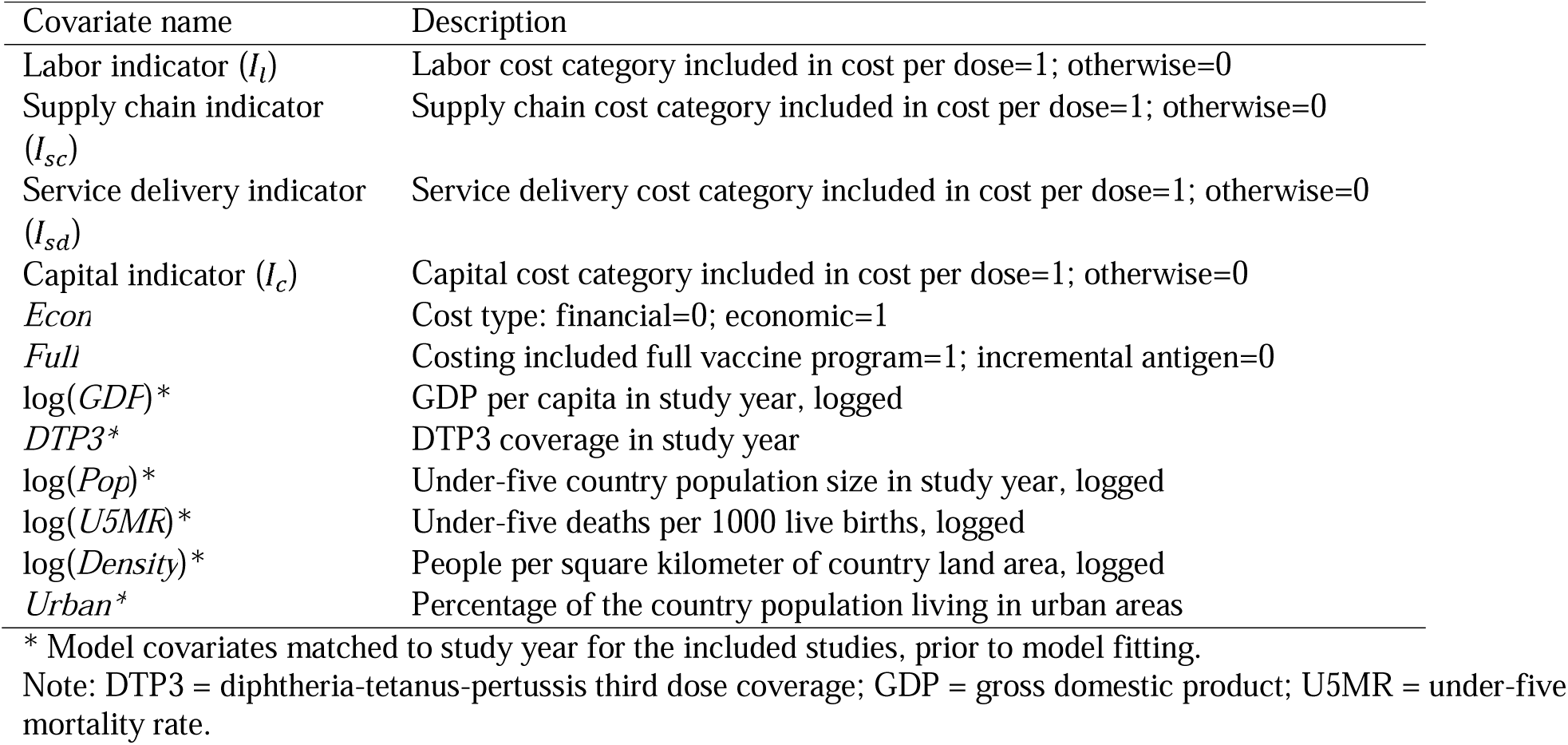
Model covariates, prior to model fitting.

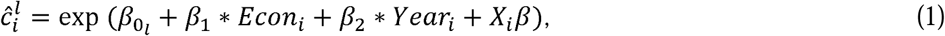

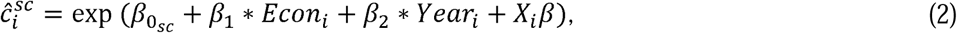

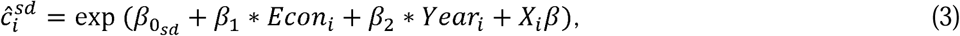

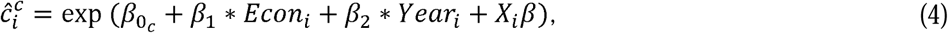

Where *X*_i_*β* represents the set of covariates selected for inclusion based on model fit for routine childhood, routine adolescent, and campaign vaccine delivery for a given study *i*. Each regression equation included indicators for economic or financial costs (*Econ_i_*) and a covariate for study year(*year_i_*). This set of equations was estimated separately for full costing studies and incremental costing studies. An additional equation related these cost category unit costs to the combination of cost categories included in each empirical study estimate, with *tc_i_* orepresenting the mean estimate of the delivery cost per dose for a given study *i*:

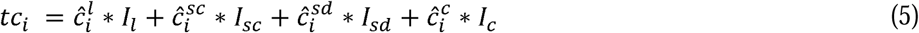

We constructed a prediction model for the intervention cost per dose outcome, specified as a generalized linear regression model (GLM), assuming a Gamma distributed outcome and a log link function. In addition to study-level predictors, we selected country-level covariates according to the best fit model using minimized Watanabe-Akaike Information Criterion (WAIC), and leave-one-out cross validation (LOOCV) as a robustness check. The model specification assumed a Gamma likelihood function for the observed data y*_i_* where the shape parameter *α* described the residual variance:

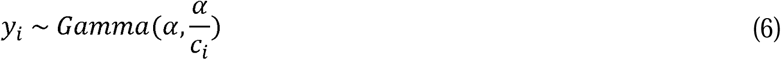

This specification assumed variance proportional to *c_i_*, as recommended for positive, right-skewed cost data [30]. We did not adjust this likelihood to account for the number of cost categories included in a given observation, assuming the likelihood reflects measurement error and unmodelled variation at the level of the observation, not the individual cost category. We assumed informative prior distributions for all regression coefficients, which were assumed to follow a normal distribution centered at zero with a standard deviation of 0.5 [31]. This prior was chosen by comparing models using Normal(0,1) and Normal(0,0.5) coefficient priors based on expected log pointwise predictive density (ELPD) estimated using leave-one-out cross validation; the Normal(0,0.5) specification yielded higher ELPD and was therefore used in the final models. The shape parameter was assumed to follow a half-Cauchy distribution centered at zero with a standard deviation of 5 [32]. To account for clustering of observations within countries and residual between-country heterogeneity not explained by the included covariates, we included country-level random intercepts. These random effects were assumed to follow a normal distribution centered at zero, with the standard deviation assigned a half-Normal(0,0.5) prior.

The prediction model was estimated in R software, version 4.4.1, using an adaptive Hamiltonian Monte Carlo algorithm in the Stan software package, version 2.32.6, with four chains of 5000 iterations. The first 2500 iterations were discarded, yielding 10,000 posterior draws for analysis [33]. Model diagnostics were examined to determine any problems encountered by the sampler, and the potential scale reduction factor (i.e., Rhat) was evaluated for all parameters to confirm that the model had successfully converged.

The fitted prediction model was used to generate both economic and financial full cost and incremental cost per dose estimates for each LMIC for the year 2024. To reduce extrapolation beyond the range of country characteristics represented in the empirical costing data, continuous country-level covariates used for prediction were constrained to the observed range of the data used to fit each model. For each model, prediction values below the observed minimum were set to the minimum and values above the observed maximum were set to the maximum before transformation and prediction. This constraint was applied separately for each delivery modality and costing approach. To generate these estimates, we predicted values from the posterior predictive distribution of the fitted model using covariate values specific to each country and year. For countries represented in the empirical costing data used to fit a given model, predictions incorporated the posterior distribution of the estimated country-specific random effect. For countries without empirical costing observations, a country random effect was drawn from the country random effects distribution estimated for each posterior draw. Therefore, prediction intervals incorporated uncertainty in the model parameters as well as residual between-country heterogeneity for countries without empirical costing data. Global cost per dose estimates were calculated as averages weighted by the estimated under-five population of individual countries for childhood vaccination and by the estimated 10–14-year-old population of individual countries for adolescent vaccination and further stratified by World Bank income level for fiscal year 2026 [21] or WHO region to produce income-level and regional cost per dose estimates, respectively. Population-weighted estimates were calculated separately for each posterior predictive draw, incorporating both parameter uncertainty and between-country heterogeneity through the aggregated estimates. For countries represented in the empirical costing data, predictions incorporated the posterior distribution of the estimated country-specific random effect; for countries without empirical costing observations, country-level random effects were drawn from the estimated random effects distribution. Reported 95% prediction intervals were defined by the 2.5^th^ and 97.5^th^ percentiles of these posterior predictive distributions. We tested predictive performance by comparing model predictions to the observed cost per dose from the IDCC 2024 update [16] matched to country and year of observation for the covariate values. We additionally assessed model fit by calculating the mean absolute error (MAE) between observed costs and posterior mean fitted costs for each final model. As a sensitivity analysis, we re-estimated the campaign delivery cost models after excluding observations for oral cholera vaccine (OCV) campaigns to assess the extent to which OCV costing studies influenced the campaign cost estimates.

## 3. Results

### 3.1 Data

We identified 32 studies across 20 countries with delivery costs for routine childhood vaccines reported in the IDCC published between 2001 and 2022, from which 141 observations (66 economic costs and 75 financial costs; 78 full costing and 63 incremental costing) were extracted and included in the analysis (Table 2). For routine HPV vaccination delivery, we identified 12 studies across 15 countries, from which 49 observations (18 economic costs and 31 financial costs; 29 full costing and 20 incremental costing; 24 school-based delivery, 6 facility-based delivery, 2 outreach/mobile delivery, and 17 assessing multiple delivery strategies) were extracted and included in the analysis. For campaign vaccination delivery, we identified 34 studies across 23 countries, from which 104 observations (52 economic costs and 52 financial costs; 76 full costing and 28 incremental costing) were extracted and included in the analysis. The majority of campaign studies represented single-antigen vaccine delivery, but 23% of the observations included integrated delivery of multiple vaccines or multiple vaccines alongside vitamin A supplementation and albendazole provision. Across the empirical studies, the observed economic full cost per dose in 2024 USD ranged from $0.21 to $21.98 (mean: $3.83), $1.52 to $20.51 (mean: $8.58), and $0.03 to $10.50 (mean: $2.25) for routine childhood, routine HPV, and campaign vaccine delivery, respectively. The observed economic incremental cost per dose ranged from $0.08 to $13.15 (mean: $3.08), $2.81 to $15.71 (mean: $5.66), and $1.18 to $8.37 (mean: $3.29) for routine childhood, routine HPV, and campaign vaccine delivery, respectively. Across the empirical studies, the observed financial full cost per dose in 2024 USD ranged from $0.21 to $36.24 (mean: $2.54), $0.46 to $7.61 (mean: $2.03), and $0.02 to $5.88 (mean: $1.13) for routine childhood, routine HPV, and campaign vaccine delivery, respectively. The observed financial incremental cost per dose ranged from $0.10 to $9.36 (mean: $1.93), $2.24 to $8.52 (mean: $3.25), and $0.18 to $6.72 (mean: $1.77) for routine childhood, routine HPV, and campaign vaccine delivery, respectively.

**Table 2.** Summary characteristics for unit cost per dose observed data.

| Variable | Routine childhood vaccine delivery | Routine adolescent vaccine delivery | Campaign vaccine delivery |
| --- | --- | --- | --- |
|  | *n* of 141 | *n* of 49 | *n* of 104 |
| Income level<sup>a</sup> |  |  |  |
| • Low | 21 | 13 | 51 |
| • Lower Middle | 77 | 31 | 51 |
| • Upper Middle | 43 | 3 | 2 |
| World region |  |  |  |
| • AFR | 81 | 25 | 69 |
| • AMR | 2 | 5 | 5 |
| • EMR | 1 | 0 | 0 |
| • EUR | 11 | 0 | 0 |
| • SEAR | 43 | 7 | 18 |
| • WPR | 3 | 12 | 12 |
| Cost type<sup>b</sup> |  |  |  |
| • Economic | 66 | 18 | 52 |
| • Financial | 75 | 31 | 52 |
| Costing approach<sup>c</sup> |  |  |  |
| • Full | 78 | 29 | 76 |
| • Incremental | 63 | 20 | 28 |
| **Average (Minimum–Maximum) across observed data** |  |  |  |
| Per-capita GDP | \$1970 (\$305–8016) | \$1756 (\$565–6406) | \$1560 (\$400–7983) |
| Under-five population (thousands) | 18532 (59–119616) | 8283 (64–129882) | 17215 (1115–128478) |
| DTP3 coverage | 89% (73–99%) | 90% (68–99%) | 84% (53–98%) |
| Under-five mortality rate | 46 (11–154) | 51 (7–127) | 70 (21–137) |
| Population density | 139 (18–476) | 146 (4–518) | 259 (15–1301) |
| Percentage urban population | 39% (17–71%) | 30% (16–76%) | 39% (16–75%) |

<sup>a</sup> World Bank income levels for fiscal year 2026 [21].
<sup>b</sup> Financial costs refer to accounting transactions (i.e., expenditures or monetary outlays) whereas economic costs include both financial costs as well as the value of resources based on their opportunity cost, regardless of whether a financial transaction occurred (e.g., community outreach workers' volunteer labor).
<sup>c</sup> Full costing represents the ongoing (i.e., recurrent) costs associated with vaccine delivery whereas incremental costing represents both the start-up (i.e., introduction) and delivery costs associated with adding a vaccine to the national immunization program.
Note: AFR = African region; AMR = Region of the Americas; DTP3 = diphtheria-tetanus-pertussis third dose; EMR = Eastern Mediterranean region; EUR = European region; GDP = gross domestic product; SEAR = Southeast Asian region; WPR = Western Pacific region.

### 3.2 Regression model

Table 3 reports posterior means and standard deviations for regression coefficients and other model parameters. For routine childhood full delivery costs, the best fit model specification according to minimized WAIC included covariates log(*GDP*) and log(*POP*). For routine adolescent vaccine full delivery costs, the best fit model specification according to minimized WAIC included covariates log(*Density*). For campaign vaccine full delivery costs, the best fit model specification according to minimized WAIC included covariates log(*GDP*), log(*POP*), *DTP3*, log(*U5MR*) and log(*Density*). For routine childhood incremental delivery costs, the best fit model specification according to minimized WAIC included covariates log(*GDP*), log(*POP*), *DTP3*, and log(*U5MR*). For routine adolescent vaccine incremental delivery costs, the best fit model specification according to minimized WAIC included covariates *DTP3* and log(*U5MR*). For campaign vaccine incremental delivery costs, the best fit model specification according to minimized WAIC included covariates log(*POP*), *DTP3*, log(*U5MR*) and log(*Density*). Appendix Table B provides an assessment of WAIC and LOOCV across models. Both statistics corroborated the best fit model. Figure 1 displays the in-sample fit comparing observed [16] versus predicted values for the study sample for each delivery modality and costing methodology. Across the six final models, MAE between observed and posterior mean fitted costs ranged from $0.49 to $2.11 per dose, with MAEs of $2.11 and $1.33 for routine childhood full and incremental costs, $1.57 and $0.79 for routine adolescent costs, and $0.49 and $0.59 for campaign costs, respectively. Residual between-country heterogeneity remained after accounting for the included study-and country-level covariates. The estimated between-country standard deviation of the random intercept was 0.33 for routine childhood full costs and 0.20 for incremental costs. Corresponding estimates were 0.45 and 0.27 for routine adolescent delivery and 1.47 and 0.57 for campaign delivery, respectively. Between-country heterogeneity was therefore greatest for full campaign delivery costs.

**Figure 1.**
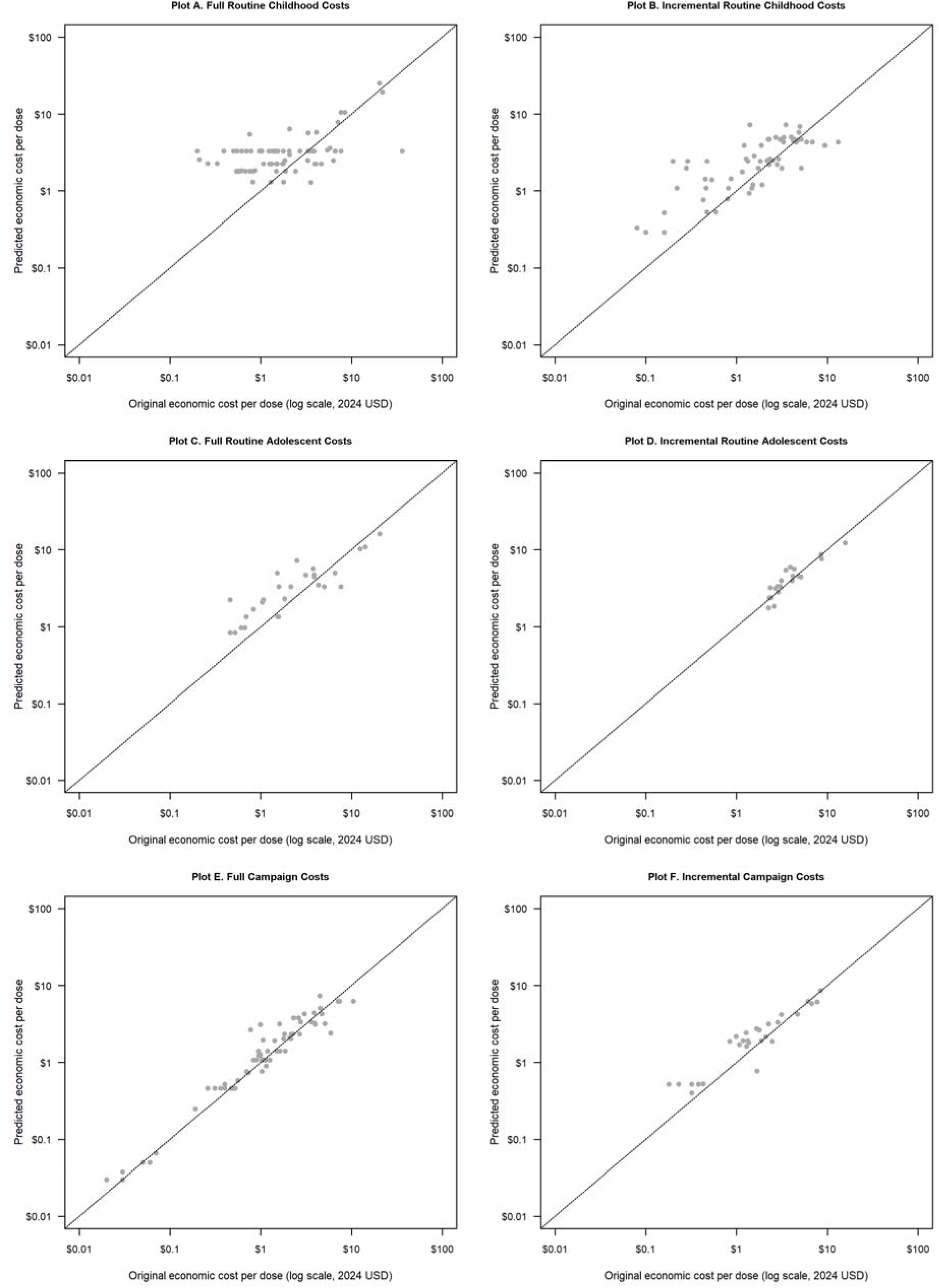
Comparison of predicted economic cost per dose and published literature economic cost per dose [16].

**Table 3.** Mean coefficient and standard errors for regressions of delivery unit cost per dose on predictors by delivery modality and costing methodology.

| Variable | Routine childhood |  | Routine adolescent |  | Campaign |  |
| --- | --- | --- | --- | --- | --- | --- |
|  | Full | Incremental | Full | Incremental | Full | Incremental |
| **Cost category intercepts** |  |  |  |  |  |  |
| Labor | -0.2898 (0.3989) | -0.6721 (0.3549) | -0.3222 (0.3991) | 0.4555 (0.2262) | -0.6552 (0.4181) | -0.5186 (0.3837) |
| Supply chain | -0.2929 (0.3973) | -0.7791 (0.3510) | -0.3183 (0.3981) | -0.0850 (0.3910) | -0.6539 (0.4099) | -0.5175 (0.3896) |
| Service delivery | -0.4102 (0.4043) | -0.7310 (0.3563) | -0.3246 (0.3993) | -0.0762 (0.3895) | -0.4883 (0.4136) | -0.5241 (0.3810) |
| Capital | -0.3769 (0.4076) | -0.1471 (0.4096) | 0.0029 (0.5024) | -0.0009 (0.4867) | -0.4002 (0.3766) | -0.1408 (0.4791) |
| **Predictors** |  |  |  |  |  |  |
| Economic cost indicator | 0.2321 (0.2500) | 0.6006 (0.1882) | 0.8271 (0.3146) | 0.3409 (0.1307) | 0.5062 (0.0983) | 0.3826 (0.2136) |
| Year | -0.1015 (0.1215) | 0.4201 (0.1488) | 0.4289 (0.1896) | 0.3116 (0.1203) | -0.1916 (0.1296) | -0.1564 (0.2183) |
| log(GDP) | 0.4156 (0.1642) | 0.3838 (0.1497) | Excluded* | Excluded* | -0.2489 (0.2064) | Excluded* |
| log(Pop) | -0.3095 (0.1601) | -0.1337 (0.1463) | Excluded* | Excluded* | 0.0686 (0.3265) | -0.2235 (0.2613) |
| DTP3 | Excluded* | -0.1078 (0.1943) | Excluded* | -0.2157 (0.1446) | 0.0876 (0.1447) | 0.1987 (0.2318) |
| log(U5MR) | Excluded* | -0.4718 (0.2107) | Excluded* | -0.1090 (0.1502) | -0.6177 (0.2815) | -0.0783 (0.2430) |
| log(Density) | Excluded* | Excluded* | -0.2659 (0.1816) | Excluded* | -0.1840 (0.3351) | -0.5154 (0.2029) |
| Urban | Excluded* | Excluded* | Excluded* | Excluded* | Excluded* | Excluded* |
| **Gamma dispersion parameter** |  |  |  |  |  |  |
| Alpha | 1.2440 (0.1552) | 2.0412 (0.3655) | 2.8202 (1.0430) | 12.7689 (5.6355) | 8.0560 (1.5555) | 4.3659 (1.6450) |
| **Country-level random effect standard deviation** |  |  |  |  |  |  |
| Sigma<sub>country</sub> | 0.3295 (0.2326) | 0.1976 (0.1603) | 0.4471 (0.2106) | 0.2728 (0.1619) | 1.4666 (0.2246) | 0.5715 (0.2317) |

\*Covariate excluded as part of the selection of best fit model for this cost outcome.
Note: Continuous predictors were standardized to mean zero and unit standard deviation; thus, fitted coefficients for continuous variables (e.g., log(GDP)) represent the increase in log cost per dose observed for a 1.0 standard deviation increase in the variable. Values in parentheses represent standard errors. DTP3 = diphtheria-tetanus-pertussis third dose coverage; GDP = gross domestic product per capita; Pop = under-five population; U5MR = under-five mortality rate.

### 3.3 Estimated costs per dose for all LMICs

For the year 2024, the population-weighted average economic full cost per dose for routine delivery of childhood vaccines was estimated to be $4.27 (95% prediction interval: $1.95–11.58) across all 136 LMICs (Appendix Table C), whereas the population-weighted average financial full cost per dose was estimated to be $3.42 ($1.58–8.33). The population-weighted average economic full cost per dose for routine delivery of HPV vaccines was estimated to be $20.56 ($8.56–57.83), and the equivalent financial full cost per dose was estimated to be $9.00 ($3.54– 25.24). Finally, for vaccines delivered via mass vaccination campaign, the population-weighted average economic full cost per dose was estimated to be $14.71 ($4.90–86.98) and the population-weighted average financial full cost per dose was estimated to be $8.73 ($2.90– 47.58). Excluding OCV costing observations from the campaign costing analysis resulted in a population-weighted average economic full cost per dose estimate of $14.71 ($4.90–86.98) and financial full cost per dose estimate of $8.73 ($2.90–47.58). The set of country-specific estimates for delivery full cost per dose can be found in Appendix Table D.

For the year 2024, the population-weighted average economic incremental cost per dose for routine delivery of childhood vaccines was estimated to be $16.64 ($7.90–38.22) across all 136 LMICs, whereas the population-weighted average financial incremental cost per dose was estimated to be $9.14 ($4.65–19.80). The corresponding economic and financial incremental costs per dose for routine delivery of HPV vaccines were $20.43 ($8.27–47.78) and $14.54 ($5.88–33.48), respectively. Incremental costs were substantially lower for campaign vaccination, at $3.53 ($1.70–8.56) for economic costs and $2.42 ($1.23–5.48) for financial costs. Excluding OCV costing observations from the campaign costing analysis resulted in a population-weighted average economic incremental cost per dose estimate of $4.54 ($1.88– 17.55) and financial incremental cost per dose estimate of $2.78 ($1.23–9.75). The set of country-specific estimates for delivery incremental cost per dose can be found in Appendix Table E.

Estimates also varied by World Bank income level, although patterns differed by delivery modality and costing approach. Economic full costs for routine childhood vaccination were higher in upper-middle income countries than in low- and lower-middle income countries, while routine adolescent full costs were more similar across income groups. Campaign full costs showed the opposite pattern, with the highest estimate among low-income countries. Incremental costs showed greater variation across income groups, particularly for routine childhood vaccination, while campaign incremental costs remained comparatively low across all three income groups. Complete economic and financial estimates by income level are reported in Appendix Table C. Figure 2 presents the country-level cost per dose estimates by GDP per capita and World Bank income level for 136 LMICs for each outcome.

**Figure 2.**
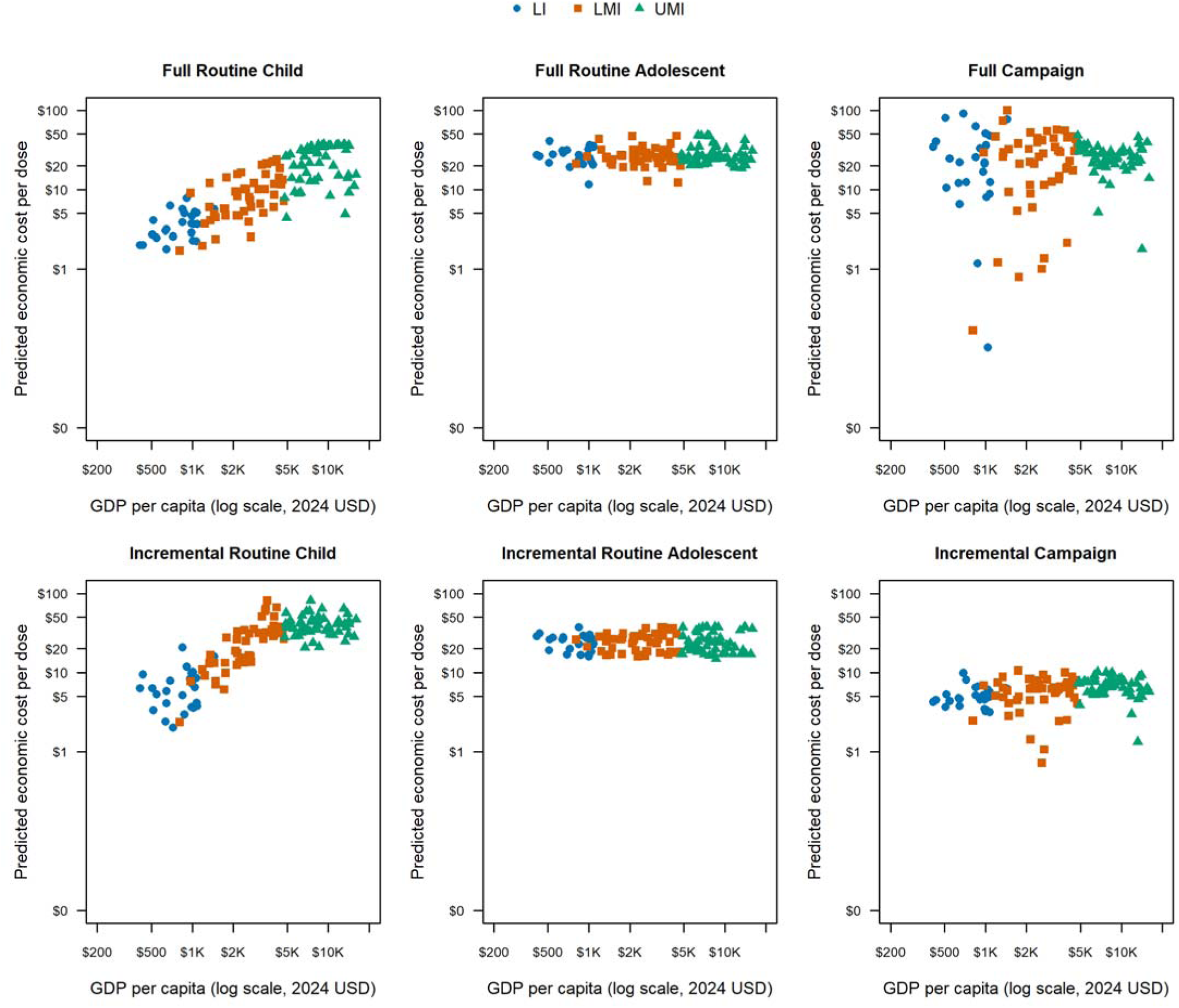
Predicted economic cost per dose in 2024 USD by per-capita gross domestic product (GDP) and World Bank income level^a^ for 136 low- and middle-income countries by delivery modality and costing methodology. ^a^LI: Gross national income (GNI) per capita of $1,135 or less in 2024; LMI: GNI per capita $1,136 and $4,495; UMI: GNI per capita of $4,496 and $13,935 (World Bank 2024) [21].

Population-weighted estimates similarly varied across WHO regions, with no single region consistently having the highest or lowest costs across delivery modalities and costing approaches. Regional variation was particularly pronounced for routine childhood vaccination, whereas campaign delivery costs were more similar across regions. Complete economic and financial full and incremental cost estimates for all six WHO regions are reported in Appendix Table C.

## 4. Discussion

Model-based estimation approaches, such as Bayesian meta-regression, can support decision-making, particularly for countries lacking recent empirical data. Using observed costs per dose from the 2024 update of the IDCC, our study provides updated delivery cost estimates for 2024 and adds routine adolescent (using HPV vaccine as a proxy) and campaign vaccine delivery to our previous routine childhood delivery estimates.

The updated empirical evidence and revised modeling approach, including separate full and incremental cost models, country-level random effects, and improved model selection and regularization, yielded substantially different estimates from our previous analysis [20]. Inflating our earlier delivery cost estimates for economic costs of routine childhood vaccine delivery from 2018 USD to 2024 USD, our meta-regression relying on the IDCC 2019 update would be equivalent to $2.41 ($0.83–5.65) per dose compared to $4.27 ($1.95–11.58) for the full economic costs of routine childhood vaccination and $16.64 ($7.90–38.22) for the incremental costs of routine childhood vaccination in the current analysis.

Given that this analysis presents both full and incremental cost per dose estimates, it is important to distinguish their respective applications. Incremental cost estimates are most relevant for short-to medium-term planning decisions, such as the introduction of a new vaccine or the scale-up of coverage. Importantly, “incremental” does not refer only to the marginal recurrent cost of delivering an additional dose. Rather, these estimates capture the additional resources required relative to an existing immunization program, including both introduction or start-up activities and additional vaccine delivery costs, and are therefore relevant to Gavi applications, co-financing projections, and investment cases. Incremental costs may consequently exceed estimates of full recurrent delivery costs, particularly when substantial introduction activities are incurred over a relatively short period. In contrast, full cost estimates reflect the total, average cost of delivering immunization services, inclusive of both fixed and variable inputs, and are more appropriate for longer-term system-level planning. These estimates can inform comprehensive resource needs assessments, medium-term expenditure frameworks, and evaluations of overall program sustainability and efficiency. However, full and incremental estimates were derived from separate subsets of the empirical costing literature and separate prediction models and therefore should not be interpreted as components of the same accounting identity or as directly comparable estimates from otherwise identical programs. For campaign delivery, full and incremental costs differ primarily in the scope of resources included. Full costs reflect broader planning and implementation requirements, whereas incremental costs may approximate the additional resources needed to conduct campaigns using existing health system capacity, excluding some shared and upstream costs.

Because the estimates are population-weighted, observations from lower middle-income countries contribute disproportionately to overall averages. While vaccine delivery costs might be expected to be lowest in low-income settings due to lower input prices, our results indicate that per-dose costs are driven by structural factors beyond price levels. Middle-income countries often vaccinate larger cohorts and achieve higher service volumes, allowing fixed logistical and programmatic costs to be spread across more doses, and may benefit from stronger health system integration and more efficient supply chains. In contrast, many low-income settings rely more heavily on outreach delivery and face greater geographic dispersion, weaker infrastructure, and smaller service volumes, all of which may increase resource requirements per dose. For routine adolescent delivery (proxied by HPV vaccination), interpretation is further limited by the concentration of observations in AFR and WPR, whose covariate profiles differ from other LMICs.

The models also indicate residual between-country heterogeneity in delivery costs beyond that explained by the observed covariates, with the greatest heterogeneity estimated for full campaign delivery costs. This suggests that measured country characteristics explain only part of the cross-country variation in campaign delivery costs. This is particularly relevant for the intended application of these estimates, as empirical delivery costing data are unavailable for many LMICs. For countries without empirical observations, the prediction intervals incorporate the estimated distribution of residual between-country heterogeneity rather than assuming that each country lies exactly at the mean conditional on its observed covariates. Accordingly, country-level point estimates should be interpreted as model-based benchmarks rather than precise estimates of local delivery costs, and the prediction intervals provide important context for their interpretation. Although these intervals reflect parameter uncertainty and residual between-country heterogeneity under the fitted model, they may not fully capture systematic differences between countries or unmeasured factors influencing delivery costs. When applying these estimates for planning, budgeting, or economic evaluation, users should therefore consider the range of predicted costs and, where possible, assess the sensitivity of their conclusions to alternative cost assumptions or locally available evidence.

The meta-regression results (Table 3) highlight associations between study-and country-level characteristics and delivery costs, with economic costs consistently exceeding financial costs in the fitted models, reflecting the inclusion of opportunity costs. For routine childhood immunization, the results are broadly consistent with expectations regarding scale and system performance, though differences emerge between full and incremental cost models. In both models, higher GDP per capita is associated with higher delivery costs, while larger population size is associated with lower costs, suggesting economies of scale. In the incremental cost model, higher DTP3 coverage and lower under-five mortality are associated with lower costs per dose, potentially reflecting greater efficiency in more established immunization systems.

For routine adolescent delivery (proxied by HPV vaccination), higher population density is associated with lower full unit costs, suggesting that geographically concentrated adolescent populations reduce logistical complexity and enable more efficient delivery, particularly for school-based strategies. In contrast, higher DTP3 coverage and lower under-five mortality are associated with lower incremental costs, potentially reflecting settings in which existing health system capacity reduces the additional resources required to expand adolescent vaccination coverage. The absence of overlapping predictors between the two models suggests that full costs primarily capture the operational realities of delivering vaccination to defined cohorts, while incremental costs reflect the marginal effort of extending coverage within existing systems. Taken together, these findings underscore that adolescent vaccination programs differ distinctly from routine childhood or campaign delivery paradigms, with total costs shaped by delivery strategy and geographic context, and marginal costs more closely tied to underlying health system capacity. However, given the heterogeneity in delivery strategies and reliance on HPV vaccination as a proxy, these interpretations should be made cautiously.

Campaign delivery exhibits the strongest associations with predictors, though these differ between full and incremental specifications. In the full cost model, costs are higher in settings with lower GDP per capita, smaller populations, lower population density, and higher under-five mortality—consistent with the expectation that campaigns are more resource-intensive in settings with weaker infrastructure and greater logistical challenges. In the incremental cost model, some of these relationships differ in direction or magnitude (e.g., population size and under-five mortality), potentially reflecting differences in the resources captured by the two costing approaches. Although predicted campaign delivery costs are lower than routine delivery costs in some countries, this should not be interpreted as evidence that campaigns are inherently more cost-efficient. While campaigns may achieve lower per-dose costs in certain settings, particularly when reaching large numbers of un- or under-immunized children, these differences may also reflect variation in delivery strategies, scale, and costing methodologies represented in the underlying studies, including incomplete capture of start-up and preparatory activities. Moreover, lower per-dose costs do not necessarily capture the broader opportunity costs of diverting resources from routine immunization and other essential health services. Because the estimates are derived from separate models and evidence bases, comparisons across delivery modalities should be interpreted cautiously and account for the specific delivery strategy and costing approach relevant to the intended application.

Across routine and campaign delivery platforms, differences in cost distributions by category highlight how cost structures vary by costing approach. For routine childhood immunization, incremental models show larger negative intercepts for labor and supply chain relative to full cost models, consistent with the exclusion of fixed and shared system inputs and reflecting economies of scale and performance-related efficiencies. In contrast, patterns for routine adolescent and campaign delivery are less consistent, indicating greater heterogeneity in cost definitions and measurement across studies. Overall, these findings emphasize that observed cost patterns differ by delivery modality and by whether studies estimated full or incremental costs. These patterns may reflect differences in the resources required for each delivery modality and the resources captured by full versus incremental costing approaches, as well as differences in underlying costing methodologies that were not fully controlled for in our models.

Beyond economic evaluation, these estimates can inform national immunization strategies, budgeting, and domestic resource mobilization [34]. Financial cost estimates are relevant to budgetary planning and fiscal space analyses, whereas economic costs, which include opportunity costs, are more appropriate for broader resource assessments and efficiency comparisons. The country-level estimates (Appendix Tables D and E) provide benchmarks for countries lacking recent empirical data, including those transitioning toward greater domestic financing responsibility. However, they should complement, rather than replace, locally generated evidence when available.

There are several limitations to this analysis, many of which are inherent to the underlying body of costing evidence. First, unobserved study characteristics may influence estimated associations between covariates and delivery costs. For example, some observed and predicted financial costs exceeded economic costs, contrary to expectations for otherwise comparable estimates. Economic and financial costs were reported by partially-overlapping sets of studies, and differences in study settings, included cost categories, and other methodological characteristics may contribute to these counterintuitive patterns. Similarly, the association between study year and cost differed in direction and magnitude across delivery modalities and costing approaches. Because the composition of the underlying evidence base changes over time with respect to countries, vaccines, delivery strategies, and study methodologies, these associations may reflect changes in the studies represented in the dataset as well as underlying changes in delivery costs.

Second, Figure 1 illustrates that predicted costs generally tracked the observed data, although predictions were more concentrated toward the center of the observed distribution. This pattern suggests regression-based shrinkage toward the mean, consistent with the regularization imposed by the model, and indicates that the covariates included in the meta-regression do not fully explain cross-study and cross-country heterogeneity. Consequently, predicted values for settings with unusually low or high delivery costs—such as remote, fragile, or small-population contexts—should be interpreted with caution. Additionally, the studies included in the dataset differed substantially in scope, costing methods, and reporting practices. Prior systematic reviews have documented similar variation in immunization costing methodologies [35–39]. Although we employed a modeling strategy designed to accommodate differences in reported cost categories and distinctions between financial and economic costs, this did not eliminate all cross-study inconsistencies. Country-level random effects account for residual heterogeneity between countries not explained by the included covariates; however, remaining unexplained variation may also reflect methodological heterogeneity across studies, including differences in costing scope, data collection, and reporting practices. All studies were conducted from the immunization provider perspective and excluded caregiver time and transportation costs; however, the operationalization of this perspective may have differed across settings. While we standardized cost categories to improve comparability, variation likely remains, particularly in the treatment of capital or investment costs (e.g., assumed useful life, discounting, and annuitization).

A third limitation is that the analysis assumes that vaccine delivery costs do not vary systematically by vaccine product. In practice, delivery costs may differ by product characteristics—for example, injectable versus oral vaccines, differences in required training or cold chain handling, or the extent to which multiple antigens are co-administered during a single visit [40, 41]. This is particularly relevant to the estimated costs of campaign vaccine delivery; twelve of 34 studies (35%) of campaign vaccine delivery describe the costs of mass vaccination with OCV, which may contribute to lower estimated costs per dose of campaign vaccination. In sensitivity analyses excluding OCV costing observations, the population-weighted economic full cost remained $14.71 per dose and the financial full cost remained $8.73 per dose; only one full-cost observation was excluded. However, for incremental costs, the economic estimate increased from $3.53 to $4.54 per dose and the financial estimate increased from $2.42 to $2.78 per dose. These changes remained within the original prediction intervals, although vaccine mix may influence campaign cost estimates.

Fourth, although this meta-regression incorporates a relatively large number of studies, most primary studies included only a limited number of service delivery sites. Estimates derived from small samples may not fully capture within-country heterogeneity, particularly where facility-level variation is substantial [42]. Although the country random effects distribution propagates residual between-country heterogeneity into predictions for countries without empirical observations, it cannot account for systematic differences between represented and unrepresented countries that are not captured by the included covariates. Moreover, the countries represented in each analytic dataset do not constitute a representative sample of all LMICs. While the regression models adjust for observable country characteristics, limited overlap between the sampled countries and the full population increases uncertainty for settings whose covariate profiles fall outside the observed range. To reduce the influence of extreme coefficient estimates when extrapolating beyond the observed data, the models used informative priors selected based on out-of-sample predictive performance. However, informative priors and country-level random effects cannot fully eliminate uncertainty when predicting costs for countries whose characteristics differ substantially from those represented in the empirical costing literature. For example, the empirical data supporting the routine childhood incremental model represented a relatively limited range of country characteristics. To reduce unsupported extrapolation when applying the model across all LMICs, country-level covariates used for prediction were constrained to the model-specific ranges represented in the empirical evidence. This approach limits extrapolation beyond the observed marginal range of individual covariates but does not ensure that all combinations of covariate values are represented in the underlying data. Moreover, for countries with characteristics outside an observed range, predictions effectively assume that the relationship between that covariate and delivery cost remains constant beyond the nearest observed boundary. Country-level estimates for settings that differ substantially from those represented in the empirical costing literature (as described in section 3.1) should therefore be interpreted cautiously. Additionally, the underlying literature search for the IDCC was also restricted to studies with full text available in English, French, or Spanish. Relevant costing studies published in other languages may therefore have been omitted, potentially contributing to uneven representation of empirical cost evidence across countries and WHO regions and increasing uncertainty in model-based predictions for underrepresented settings.

Finally, in estimating delivery costs for routine adolescent immunization, we relied on available data from HPV vaccination programs as a proxy. While HPV vaccination represents the most widely implemented routine adolescent vaccine platform in many LMICs, delivery strategies for HPV vaccination—often school-based or campaign-style approaches targeting specific age cohorts—may differ from those used for other adolescent vaccines and adult vaccines delivered through facility-based services or integrated primary care platforms (including but not limited to meningococcal conjugate vaccine and tetanus-diphtheria-pertussis booster). HPV programs in most LMICs only target females, who may be more expensive to reach on average than males, given lower school enrollment rates among girls [43–45]. Additionally, population weighting by ages 10–14 years does not fully reflect variation in HPV vaccination target ages and introduction strategies across countries.

This study cannot fully address the persistent evidence gap arising from the limited number of immunization costing studies relative to the diversity of settings and policy needs, and the modeled estimates inevitably reflect the constraints of the underlying data. While continued investment in primary cost data collection remains essential, policy decisions must often be made in the absence of country-specific evidence, relying instead on regional or global benchmarks. In this context, the estimates presented here provide a practical and transparent resource to support cost-effectiveness analyses, planning, and budgeting [34], serving as a complementary input where empirical data are unavailable, outdated, or uncertain.

## Data Availability

The data that support the findings of this study are openly available in the Immunization Delivery Cost Catalogue (IDCC) at https://immunizationeconomics.org/ican-idcc/. All data produced in the present work are contained in the manuscript and supplementary materials.

## Author contributions

AP conceptualized and designed the updated study that builds on a previous study conceptualized and designed by AP and NAM. AP did the analysis, drafted the initial manuscript, and approved the final manuscript as submitted. ECD, TAH, DRH, TM, and NAM developed the methodology, critically reviewed the analysis, reviewed and revised the manuscript, and approved the final manuscript as submitted. AOA, LB, BNP, and SYS critically reviewed the analysis, reviewed and revised the manuscript, and approved the final manuscript as submitted.

## Code availability

The programming code is available from the corresponding author on reasonable request.

## References

1. United Nations. Sustainable development goals (SDGs). Geneva: United Nations; 2015. Last updated: September 2015. [Online] Accessed 12 February 2018. Available at: http://www.un.org/sustainabledevelopment/sustainable-development-goals/. 2015.

2. Boidin B. La sante, bien public mondial ou bien commun? Medecine et maladies infectieuses. 2014;44(5):193–200.

3. Immunization Agenda 2030 Partners. Immunization agenda 2030: A global strategy to leave no one behind. Vaccine. 2024;42 Suppl 1:S5–s14. doi: 10.1016/j.vaccine.2022.11.042. PubMed PMID: 39004466.

4. Saxenian H, Alkenbrack S, Freitas Attaran M, Barcarolo J, Brenzel L, Brooks A, et al. Sustainable financing for Immunization Agenda 2030. Vaccine. 2024;42 Suppl 1:S73–s81. Epub 20221202. doi: 10.1016/j.vaccine.2022.11.037. PubMed PMID: 36464542.

5. World Health Organization. Immunization Agenda 2030: a global strategy to leave no one behind. Last updated: 1 April 2020. [Online] Accessed 6 June 2025. Available at: https://www.who.int/publications/m/item/immunization-agenda-2030-a-global-strategy-to-leave-no-one-behind.

6. Han J, Zhang L, Chen Y, Zhang Y, Wang L, Cai R, et al. Global HPV vaccination programs and coverage rates: a systematic review. EClinicalMedicine. 2025;84:103290. Epub 20250609. doi: 10.1016/j.eclinm.2025.103290. PubMed PMID: 40547442; PubMed Central PMCID: PMCPMC12179740.

7. World Health Organization. WHO-UNICEF estimates of DTP3 coverage: monitoring system 2024 global summary. Last updated: 15 Jul 2025. [Online] Accessed 18 Sep 2025. Available at: https://immunizationdata.who.int/global

8. World Health Organization. The Big Catch-Up: An Essential Immunization Recovery Plan for 2023 and Beyond. Last Updated; 26 July 2023. [Online] Accessed 6 June 2025. Available at: https://www.who.int/publications/i/item/9789240075511.

9. Hartner AM, Li X, Echeverria-Londono S, Roth J, Abbas K, Auzenbergs M, et al. Estimating the health effects of COVID-19-related immunisation disruptions in 112 countries during 2020-30: a modelling study. Lancet Glob Health. 2024;12(4):e563–e71. doi: 10.1016/s2214-109x(23)00603-4. PubMed PMID: 38485425; PubMed Central PMCID: PMCPMC10951961.

10. Kumar A, Gabani J, Marino A, Ramirez JCM, Eozenou PHV. At a Crossroads: Prospects for Government Health Financing Amidst Declining Aid Government Resources and Projections for Health (GRPH) Series Washington, DC: World Bank, November 2025 https://hdlhandlenet/10986/43745.

11. Audibert M, Mathonnat J, de Roodenbeke E. [Evolution and new perspectives of health care financing in developing countries]. Sante. 2003;13(4):209–14. PubMed PMID: 15047437.

12. Dieleman JL, Schneider MT, Haakenstad A, Singh L, Sadat N, Birger M, et al. Development assistance for health: past trends, associations, and the future of international financial flows for health. Lancet (London, England). 2016;387(10037):2536–44. Epub 20160413. doi: 10.1016/s0140-6736(16)30168-4. PubMed PMID: 27086170.

13. Kaddar M, Schmitt S, Tanaka S, Battison J. Tendances mondiales du financement national des programmes de vaccination dans les pays a revenu faible et intermediaire. Bulletin de l’Organisation mondiale de la Sante. 2013;91(9):688–95.

14. Gavi The Vaccine Alliance. Gavi, the Vaccine Alliance Co-Financing Policy. Version No. 4.0. Geneva: Gavi, The Vaccine Alliance. Last updated: 2 July 2026. [Online] Accessed 28 September 2026. Available at: https://www.gavi.org/about/programme-policies/co-financing/. 2026.

15. Mao W, McDade KK, Ogbuoji O, Yamey G, Bermeo SB. Strategic donor behaviour and country vulnerability in health aid transitions. BMJ Glob Health. 2023;8(11). doi: 10.1136/bmjgh-2023-012953. PubMed PMID: 37940202; PubMed Central PMCID: PMCPMC10632813.

16. Immunization Costing Action Network (ICAN). Immunization delivery cost catalogue (IDCC). ThinkWell. Last updated: 20 Jun 2024. [Online] Accessed 28 Sep 2024. Available at: https://immunizationeconomics.org/thinkwell-idcc/

17. UNICEF. Costs of Fully Vaccinating a Child. August 2024. [Online] Accessed 25 September 2025. Available at: https://www.unicef.org/documents/costs-fully-vaccinating-child.

18. Dixit S, Diab MM, Zimmerman A, Ogbuoji O, Yamey G. COVID-19 vaccination cost map. The Center for Policy Impact in Global Health. Dialogue report: January 2021. Available at: https://centerforpolicyimpact.org/our-work/covid-19-vaccine-cost-map/.

19. World Health Organization. An investment case for new tuberculosis vaccines. Geneva: World Health Organization; 2022. Licence: CC BY-NC-SA 3.0 IGO. https://www.who.int/publications/i/item/9789240064690.

20. Portnoy A, Vaughan K, Clarke-Deelder E, Suharlim C, Resch SC, Brenzel L, et al. Producing Standardized Country-Level Immunization Delivery Unit Cost Estimates. Pharmacoeconomics. 2020;38(9):995–1005. doi: 10.1007/s40273-020-00930-6. PubMed PMID: 32596785; PubMed Central PMCID: PMCPMC7437655.

21. World Bank. World Bank Country and Lending Groups. Washington, DC: The World Bank. Last updated: 1 July 2025. [Online] Accessed 18 Sep 2025. Available at: https://datahelpdesk.worldbank.org/knowledgebase/articles/906519.

22. Immunization Costing Action Network (ICAN). Immunization delivery cost catalogue (IDCC). ThinkWell. Last updated: 20 Jun 2024. [Online] Accessed 20 Jun 2024. Available at: https://immunizationeconomics.org/thinkwell-idcc/

23. Vaughan K, Ozaltin A, Mallow M, Moi F, Wilkason C, Stone J, et al. The costs of delivering vaccines in low-and middle-income countries: Findings from a systematic review. Vaccine X. 2019;2:100034. Epub 20190715. doi: 10.1016/j.jvacx.2019.100034. PubMed PMID: 31428741; PubMed Central PMCID: PMCPMC6697256.

24. Brenzel L, Young D, Walker DG. Costs and financing of routine immunization: Approach and selected findings of a multi-country study (EPIC). Vaccine. 2015;33 Suppl 1:A13–20. Epub 2015/04/29. PubMed PMID: 25919153.

25. Resch S, Menzies NA, Portnoy A, Clarke-Deelder E, O’Keeffe L, Suharlim C, et al. HOW TO COST IMMUNIZATION PROGRAMS: A Practical Guide on Primary Data Collection and Analysis. December 2020. [Online] Accessed 17 Feb 2026. Available at: https://immunizationeconomics.org/recent-activity/2020/12/23/how-to-cost-guide/.

26. International Monetary Fund. World economic outlook database. April 2025. [Online] Accessed 24 Sept 2025. Available at: https://www.imf.org/en/publications/weo/weo-database/2025/april.

27. World Bank. World development indicators. Last updated: 1 Jul 2025. [Online] Accessed 18 Sep 2025. Available at: https://data.worldbank.org/

28. World Bank. World development indicators. Washington, DC: The World Bank; 2019. Last updated: 10 July 2019. [Online] Accessed 13 August 2019. Available at: http://data.worldbank.org/. 2019.

29. World Health Organization. WHO-UNICEF estimates of DTP3 coverage: monitoring system 2019 global summary. Geneva: WHO/UNICEF. Last updated: 15 July 2019. [Online] Accessed 13 August 2019. Available at: http://apps.who.int/immunization_monitoring/globalsummary/timeseries/tswucoveragedt p3.html. 2019.

30. Manning WG, Mullahy J. Estimating log models: to transform or not to transform? Journal of Health Economics. 2001;20(4):461–94.

31. Gelman A. Prior Choice Recommendations. Last updated: 27 April 2025. [Online] Accessed 25 May 2025. Available at: https://github.com/stan-dev/stan/wiki/Prior-Choice-Recommendations.

32. Gelman A. Prior distributions for variance parameters in hierarchical models. Bayesian Analysis. 2006;1(3):515–34. doi: 10.1214/06-BA117A.

33. Hoffman MD, Gelman A. The No-U-Turn Sampler: Adaptively Setting Path Lengths in Hamiltonian Monte Carlo. Journal Of Machine Learning Research. 2014;15:1593–623.

34. Perrin de Brichambaut C, Golaz A, Abiteboul D, Jestin C. Evaluation economique de la vaccination: principes et methodes. Bulletin epidemiologique hebdomadaire. 2011;14(15):165–7.

35. Batt K, Fox-Rushby JA, Castillo-Riquelme M. The costs, effects and cost-effectiveness of strategies to increase coverage of routine immunizations in low-and middle-income countries: systematic review of the grey literature. Bull World Health Organ. 2004;82(9):689–96. Epub 2005/01/05. PubMed PMID: 15628207; PubMed Central PMCID: PMCPMC2622984.

36. Clarke-Deelder E, Vassall A, Menzies NA. Estimators Used in Multisite Healthcare Costing Studies in Low-and Middle-Income Countries: A Systematic Review and Simulation Study. Value Health. 2019;22(10):1146–53. Epub 2019/09/30. PubMed PMID: 31563257; PubMed Central PMCID: PMCPMC6859917.

37. Munk C, Portnoy A, Suharlim C, Clarke-Deelder E, Brenzel L, Resch SC, et al. Systematic review of the costs and effectiveness of interventions to increase infant vaccination coverage in low- and middle-income countries. BMC Health Serv Res. 2019;19(1):741.

38. Ozawa S, Yemeke TT, Thompson KM. Systematic review of the incremental costs of interventions that increase immunization coverage. Vaccine. 2018;36(25):3641–9. Epub 2018/05/15. PubMed PMID: 29754699.

39. Pegurri E, Fox-Rushby JA, Walker DG. The effects and costs of expanding the coverage of immunisation services in developing countries: a systematic literature review. Vaccine. 2004;23(13):1624–35. Epub 2005/02/08. PubMed PMID: 15694515.

40. Boonstoppel L, Moi F, Banks C, Sibeudu F, Obodoechi D, Borces K, et al. Cost of integrated immunization campaigns in Nigeria and Sierra Leone: bottom-up costing studies. BMC Health Serv Res. 2024;24(1):1334. Epub 20241101. doi: 10.1186/s12913-024-11809-z. PubMed PMID: 39487478; PubMed Central PMCID: PMCPMC11529071.

41. Debellut F, Bello G, Chisema M, Mkisi R, Kamzati M, Pecenka C, et al. Cost of the typhoid conjugate vaccine introduction through an integrated campaign and follow-on routine immunization in Malawi. Vaccine X. 2024;21:100583. Epub 20241113. doi: 10.1016/j.jvacx.2024.100583. PubMed PMID: 39633851; PubMed Central PMCID: PMCPMC11614825.

42. Adam T, Evans DB, Murray CJ. Econometric estimation of country-specific hospital costs. Cost Eff Resour Alloc. 2003;1(1):3. Epub 2003/05/30. PubMed PMID: 12773218; PubMed Central PMCID: PMCPMC156022.

43. Bonfert A, Wadhwa D. Tracing global trends in education: A tale of old and new gender gaps. World Bank Gender Data Portal. https://genderdata.worldbank.org/en/data-stories/a-tale-of-old-and-new-gender-gaps. 2024.

44. UNICEF USA. Equality for girls. Accessed 29 March 2026. Available at: https://www.unicefusa.org/what-unicef-does/respect-children/equality-for-girls.

45. Psaki S, McCarthy KJ, Mensch BS. Measuring gender equality in education: Lessons from trends in 43 countries. Population and Development Review. 2018;44(1):117–42.

