## Supplementary Appendix for "Standardized country-level delivery unit cost estimates for routine childhood, routine adolescent, and campaign vaccination: an updated modeling analysis"

**Running title:** Updated vaccine delivery cost estimates for childhood, adolescent, and campaign programs

Allison Portnoy^1,2^, Emma Clarke-Deelder^3,4,*^, Taylor A. Holroyd^5,*^, Daniel R. Hogan^5^, Alex Olateju Adjagba^6,7^, Logan Brenzel^8^, Bryan N. Patenaude^9^, So Yoon Sim^10^, Tewodaj Mengistu^5,*^, Nicolas A. Menzies^2,11,*^

* *authors contributed equally.*

**Affiliations**: ^1^Department of Global Health, Boston University School of Public Health; ^2^Center for Health Decision Science, Harvard T.H. Chan School of Public Health; ^3^Department of Research, Swiss Tropical & Public Health Institute; ^4^University of Basel; ^5^Gavi, the Vaccine Alliance; ^6^University of Western Cape, Cape Town, South Africa; ^7^Global Health Practice, UNICEF Center of Excellence for Child Survival and Development, Nairobi, Kenya; ^8^Health Economist, Paris, France; ^9^Department of International Health, Johns Hopkins Bloomberg School of Public Health; ^10^Department of Immunization, Vaccines, and Biologicals, World Health Organization, Geneva, Switzerland; ^11^Department of Global Health and Population, Harvard T.H. Chan School of Public Health.

**Appendix Table A. Immunization Delivery Cost Catalogue (IDCC) cost categories mapped to analytic cost categories.**

| **Analytic cost categories** | **IDCC cost categories** |
| --- | --- |
| Labor | Paid human resources; volunteer human resources; per diem and travel allowances |
| Service delivery | Training and capacity building; social mobilization and advocacy; program management; disease surveillance; management of adverse events following immunization |
| Supply chain | Cold chain equipment and their overheads (installation, energy, maintenance, repairs); vehicles, transport and fuel |
| Capital | Buildings, utilities, other overheads and/or shared costs |

**Appendix Table B. Model fit statistics according to Watanabe-Akaike Information Criterion (WAIC) and leave-one-out cross validation (LOOCV).**

| **Model** | **WAIC** | | | | | | **LOOCV** | | | | | |
| --- | --- | --- | --- | --- | --- | --- | --- | --- | --- | --- | --- | --- |
|  | **Routine childhood** | | **Routine adolescent** | | **Campaign** | | **Routine childhood** | | **Routine adolescent** | | **Campaign** | |
|  | **Full** | **Incremental** | **Full** | **Incremental** | **Full** | **Incremental** | **Full** | **Incremental** | **Full** | **Incremental** | **Full** | **Incremental** |
| **1** | 320.513 | 217.847 | 126.739 | 66.429 | 236.204 | 110.324 | 321.820 | 218.258 | 127.445 | 67.726 | 236.147 | 109.365 |
| **2** | **318.837** | 214.311 | 127.922 | 68.475 | 230.529 | 97.765 | **320.764** | 214.626 | 128.678 | 70.508 | 231.190 | 96.616 |
| **3** | 319.307 | 214.651 | 129.891 | 68.463 | 231.613 | 98.462 | 321.490 | 215.488 | 130.927 | 69.938 | 232.078 | 97.652 |
| **4** | 321.064 | **202.462** | 127.878 | 66.290 | 205.272 | 99.569 | 322.748 | **202.965** | 128.968 | 68.411 | 205.467 | 100.259 |
| **5** | 321.573 | 204.524 | 126.716 | 68.712 | **155.192** | 77.303 | 322.134 | 205.074 | 128.470 | 70.687 | **155.500** | 71.604 |
| **6** | 338.432 | 247.259 | 126.657 | 70.063 | 231.287 | 95.393 | 339.309 | 247.963 | 127.110 | 71.164 | 231.260 | 93.549 |
| **7** | 340.166 | 241.324 | 128.817 | 70.726 | 230.002 | 96.085 | 340.904 | 241.386 | 129.567 | 71.994 | 230.510 | 94.901 |
| **8** | 321.893 | 210.817 | 125.618 | 64.459 | 224.337 | 97.285 | 323.488 | 211.453 | 126.377 | 66.179 | 224.514 | 96.677 |
| **9** | 323.627 | 207.612 | 123.774 | 65.922 | 201.843 | **74.843** | 324.621 | 208.479 | 124.686 | 67.444 | 202.779 | **68.737** |
| **10** | 323.591 | 208.211 | 125.698 | 67.166 | 183.098 | 75.791 | 325.656 | 208.848 | 127.174 | 69.649 | 184.195 | 71.585 |
| **11** | 343.385 | 239.250 | 127.221 | 68.747 | 229.326 | 108.958 | 343.827 | 239.274 | 127.695 | 69.443 | 229.573 | 108.442 |
| **12** | 322.407 | 212.107 | 123.842 | **61.674** | 223.368 | 107.121 | 323.465 | 212.763 | 124.480 | **62.825** | 223.572 | 107.113 |
| **13** | 322.068 | 204.804 | 123.743 | 63.716 | 199.956 | 82.997 | 322.639 | 205.982 | 124.553 | 65.096 | 200.344 | 80.107 |
| **14** | 323.517 | 205.504 | 125.732 | 64.367 | 187.591 | 85.419 | 326.126 | 205.989 | 127.176 | 66.153 | 188.947 | 83.076 |
| **15** | 325.368 | 212.782 | 123.263 | 67.875 | 230.296 | 108.636 | 326.907 | 213.033 | 123.598 | 69.090 | 230.770 | 108.313 |
| **16** | 324.242 | 207.444 | 122.055 | 68.618 | 196.368 | 81.120 | 330.184 | 206.772 | 122.673 | 69.891 | 197.153 | 78.541 |
| **17** | 323.808 | 206.229 | 123.188 | 66.046 | 186.640 | 83.366 | 325.631 | 206.430 | 124.007 | 68.023 | 187.436 | 81.114 |
| **18** | 336.558 | 249.379 | **121.715** | 69.777 | 228.955 | 87.903 | 337.600 | 249.707 | **122.108** | 70.983 | 228.986 | 86.864 |
| **19** | 325.874 | 247.906 | 123.337 | 71.829 | 229.635 | 90.428 | 326.588 | 248.270 | 123.786 | 73.675 | 229.986 | 89.946 |
| **20** | 323.772 | 246.347 | 126.174 | 71.259 | 236.360 | 110.171 | 324.928 | 246.599 | 126.641 | 73.036 | 236.767 | 110.966 |
| **Best fit model** | **2** | **4** | **18** | **12** | **5** | **9** | **2** | **4** | **18** | **12** | **5** | **9** |
| **MIN** | **318.837** | **202.462** | **121.715** | **61.674** | **155.192** | **74.843** | **320.764** | **202.965** | **122.108** | **62.825** | **155.500** | **68.737** |

Note: Values in bold text represent the minimal value across models, and are listed as the best fit model.

**Appendix Table C. Predicted cost per dose in 2024 USD by delivery modality and world region/income level.**

|  | **Routine childhood** | | | **Routine adolescent** | | | **Campaign** | |
| --- | --- | --- | --- | --- | --- | --- | --- | --- |
|  | **Full** | **Incremental** | | **Full** | | **Incremental** | **Full** | **Incremental** |
| **Across LMICs** | | |  | |  | | | |
| *Economic* | 4.27 (1.95–11.58) | 16.64 (7.90–38.22) | | 20.56 (8.56–57.83) | | 20.43 (8.27–47.78) | 14.71 (4.90–86.98) | 3.53 (1.70–8.56) |
| *Financial* | 3.42 (1.58–8.33) | 9.14 (4.65–19.80) | | 9.00 (3.54–25.24) | | 14.54 (5.88–33.48) | 8.73 (2.90–47.58) | 2.42 (1.23–5.48) |
| **World region** | | |  | |  | | | |
| *Economic* |  |  | |  | |  |  |  |
| - AFR | 3.16 (1.43–8.93) | 7.87 (3.36–19.32) | | 25.24 (10.20–75.13) | | 23.34 (8.63–58.08) | 13.23 (4.24–82.23) | 3.95 (1.83–10.33) |
| - AMR | 9.56 (3.58–30.83) | 33.50 (10.81–105.87) | | 25.72 (10.07–74.75) | | 26.32 (8.68–74.17) | 9.81 (2.14–99.03) | 5.23 (2.29–13.70) |
| - EMR | 3.67 (1.68–10.61) | 13.72 (6.66–30.32) | | 21.49 (8.69–65.16) | | 18.86 (7.91–42.37) | 19.21 (4.48–170.40) | 4.07 (1.90–10.29) |
| - EUR | 10.83 (3.70–37.07) | 28.30 (12.19–68.21) | | 22.51 (8.91–67.92) | | 19.42 (7.49–46.33) | 17.77 (3.04–184.50) | 6.65 (2.94–16.01) |
| - SEAR | 2.77 (1.24–9.01) | 17.50 (8.89–37.18) | | 13.69 (5.01–40.35) | | 19.70 (7.83–46.30) | 4.34 (1.42–38.33) | 1.63 (0.83–3.63) |
| - WPR | 5.01 (2.08–14.95) | 24.53 (11.77–57.65) | | 18.16 (5.94–69.98) | | 17.24 (6.91–39.32) | 10.94 (1.56–220.86) | 3.93 (1.68–10.87) |
| *Financial* |  |  | |  | |  |  |  |
| - AFR | 2.55 (1.13–6.81) | 4.32 (1.94–10.13) | | 11.13 (4.23–31.70) | | 16.62 (6.27–40.42) | 7.98 (2.54–48.99) | 2.70 (1.30–6.70) |
| - AMR | 7.65 (2.65–23.26) | 18.34 (6.33–55.32) | | 11.37 (4.05–34.49) | | 18.67 (6.10–52.28) | 5.78 (1.24–59.41) | 3.58 (1.63–8.91) |
| - EMR | 2.95 (1.35–7.69) | 7.54 (3.97–15.52) | | 9.59 (3.45–28.39) | | 13.38 (5.62–29.68) | 11.24 (2.71–85.12) | 2.79 (1.39–6.57) |
| - EUR | 8.63 (2.62–29.29) | 15.42 (7.48–34.10) | | 10.11 (3.68–29.85) | | 13.88 (5.31–33.19) | 10.67 (1.83–104.51) | 4.56 (2.03–10.83) |
| - SEAR | 2.22 (1.08–6.00) | 9.58 (5.05–19.69) | | 6.06 (2.04–18.27) | | 14.02 (5.54–32.37) | 2.66 (0.86–24.33) | 1.11 (0.58–2.37) |
| - WPR | 4.05 (1.68–10.77) | 13.48 (6.80–29.98) | | 8.10 (2.32–29.24) | | 12.21 (4.91–28.08) | 6.20 (0.92–119.86) | 2.68 (1.26–6.93) |
| **Income level^a^** | | |  | |  | | | |
| *Economic* |  |  | |  | |  |  |  |
| - LI | 2.23 (0.94–7.73) | 5.87 (2.09–17.57) | | 22.73 (9.10–71.00) | | 22.90 (8.51–56.24) | 18.44 (5.18–116.69) | 3.67 (1.60–10.73) |
| - LMI | 3.22 (1.53–9.31) | 14.24 (7.31–29.89) | | 17.62 (7.13–49.43) | | 19.95 (8.29–45.86) | 8.56 (2.92–56.41) | 2.71 (1.34–6.34) |
| - UMI | 7.30 (2.94–21.27) | 28.46 (12.11–71.57) | | 21.37 (8.52–64.56) | | 19.95 (7.74–48.59) | 15.25 (3.08–171.09) | 4.88 (2.18–12.36) |
| *Financial* |  |  | |  | |  |  |  |
| - LI | 1.81 (0.73–5.63) | 3.21 (1.20–9.48) | | 10.10 (3.69–30.49) | | 16.33 (6.19–39.38) | 10.95 (3.10–72.22) | 2.53 (1.12–6.91) |
| - LMI | 2.60 (1.24–6.49) | 7.81 (4.22–15.43) | | 7.83 (2.94–21.82) | | 14.20 (5.88–32.22) | 5.13 (1.72–30.37) | 1.85 (0.95–4.11) |
| - UMI | 5.82 (2.30–16.03) | 15.52 (7.13–37.45) | | 9.49 (3.47–27.72) | | 14.26 (5.42–34.21) | 9.02 (1.79–95.64) | 3.32 (1.60–7.92) |

^a^ LI: Gross national income (GNI) per capita of $1,135 or less for fiscal year 2026; LMI: GNI per capita $1,136 and $4,495; UMI: GNI per capita of $4,496 and $13,935 (World Bank 2026) [18].

Note: AFR = African region; AMR = Region of the Americas; DTP3 = diphtheria-tetanus-pertussis third dose coverage; EMR = Eastern Mediterranean region; EUR = European region; GDP = gross domestic product; LI = low-income; LMI = lower middle-income; LMIC = low- and middle-income country; SEAR = Southeast Asian region; UMI = upper middle-income; WPR = Western Pacific region.

**Appendix Table D. Predicted full cost per dose in 2024 by delivery modality and country.**

| **Country** | **Income** | **Region** | **Economic Cost of Childhood Vaccine Delivery** | **Financial Cots of Childhood Vaccine Delivery** | **Economic Cost of Adolescent Vaccine Delivery** | **Financial Cost of Adolescent Vaccine Delivery** | **Economic Cost of Campaign Vaccine Delivery** | **Financial Cost of Campaign Vaccine Delivery** |
| --- | --- | --- | --- | --- | --- | --- | --- | --- |
| Afghanistan | LI | EMR | $1.37 ($0.41–7.22) | $1.10 ($0.32–4.93) | $21.03 ($5.68–89.37) | $9.49 ($2.24–35.78) | $5.67 ($0.28–124.09) | $3.50 ($0.17–78.48) |
| Albania | UMI | EUR | $24.25 ($3.76–136.93) | $19.69 ($2.52–120.30) | $19.03 ($5.22–77.99) | $8.65 ($2.11–32.08) | $4.29 ($0.17–128.22) | $2.69 ($0.11–72.52) |
| Algeria | UMI | AFR | $7.38 ($2.29–27.51) | $5.99 ($1.65–20.53) | $26.56 ($6.67–106.60) | $11.91 ($2.62–46.84) | $5.32 ($0.24–131.10) | $3.16 ($0.15–83.35) |
| Angola | LMI | AFR | $3.77 ($1.34–13.48) | $3.04 ($1.05–10.33) | $24.49 ($6.54–99.07) | $10.92 ($2.53–43.06) | $3.00 ($0.14–62.04) | $1.86 ($0.10–39.80) |
| Argentina | UMI | AMR | $10.96 ($2.84–41.60) | $8.77 ($1.99–33.65) | $27.59 ($7.03–109.04) | $12.20 ($2.61–49.73) | $3.98 ($0.18–110.41) | $2.54 ($0.10–73.76) |
| Armenia | UMI | EUR | $24.25 ($3.81–137.14) | $19.56 ($2.49–115.08) | $19.10 ($5.40–74.99) | $8.54 ($2.05–33.31) | $4.28 ($0.17–137.53) | $2.64 ($0.11–71.94) |
| Azerbaijan | UMI | EUR | $16.20 ($3.27–71.35) | $13.01 ($2.25–59.76) | $18.17 ($5.19–76.62) | $8.34 ($2.09–30.00) | $3.42 ($0.11–111.63) | $2.00 ($0.07–69.92) |
| Bangladesh | LMI | SEAR | $3.12 ($1.11–11.93) | $2.52 ($0.95–8.23) | $13.83 ($3.80–61.52) | $6.32 ($1.55–25.34) | $0.98 ($0.65–1.52) | $0.59 ($0.37–0.99) |
| Belarus | UMI | EUR | $20.15 ($3.67–99.55) | $16.19 ($2.51–84.87) | $22.48 ($6.18–88.00) | $10.03 ($2.35–39.56) | $4.63 ($0.18–142.08) | $2.78 ($0.11–80.11) |
| Belize | UMI | AMR | $24.62 ($3.74–140.77) | $19.45 ($2.57–116.11) | $27.03 ($6.86–111.36) | $12.18 ($2.64–49.55) | $4.24 ($0.16–123.71) | $2.65 ($0.09–84.07) |
| Benin | LMI | AFR | $3.77 ($0.99–11.77) | $3.05 ($0.69–9.74) | $18.46 ($5.43–76.78) | $8.25 ($2.05–32.21) | $2.35 ($0.10–51.17) | $1.44 ($0.07–31.21) |
| Bhutan | LMI | SEAR | $15.63 ($2.69–79.39) | $12.51 ($1.89–69.69) | $26.03 ($8.23–88.43) | $11.60 ($3.18–40.90) | $5.66 ($0.25–156.63) | $3.40 ($0.15–88.00) |
| Bolivia | LMI | AMR | $9.20 ($2.45–35.40) | $7.41 ($1.64–28.88) | $29.62 ($6.95–125.16) | $13.24 ($2.78–55.01) | $4.46 ($0.17–139.96) | $2.60 ($0.10–80.30) |
| Bosnia and Herzegovina | UMI | EUR | $24.38 ($3.85–132.91) | $19.56 ($2.54–120.11) | $21.16 ($5.97–81.78) | $9.50 ($2.24–35.19) | $3.96 ($0.14–116.86) | $2.49 ($0.09–74.10) |
| Botswana | UMI | AFR | $21.55 ($3.64–114.14) | $17.24 ($2.53–93.97) | $35.13 ($7.32–159.79) | $15.86 ($2.80–72.64) | $3.35 ($0.14–100.42) | $2.06 ($0.09–52.34) |
| Brazil | UMI | AMR | $6.54 ($2.03–24.69) | $5.33 ($1.55–18.28) | $25.28 ($6.65–98.22) | $11.29 ($2.62–44.24) | $4.97 ($0.22–138.65) | $3.03 ($0.15–84.73) |
| Burkina Faso | LI | AFR | $2.89 ($0.94–11.65) | $2.32 ($0.69–8.60) | $19.93 ($5.60–78.46) | $8.91 ($2.27–34.38) | $3.47 ($0.17–72.20) | $2.07 ($0.10–43.31) |
| Burundi | LI | AFR | $1.62 ($0.39–8.87) | $1.32 ($0.30–6.82) | $13.90 ($3.78–63.31) | $6.30 ($1.53–26.30) | $5.76 ($0.24–144.67) | $3.59 ($0.15–90.37) |
| Cabo Verde | UMI | AFR | $19.04 ($3.14–103.41) | $15.04 ($2.13–88.06) | $18.29 ($5.19–72.77) | $8.21 ($2.01–31.96) | $4.88 ($0.20–134.11) | $2.91 ($0.13–80.65) |
| Cambodia | LMI | WPR | $6.37 ($1.90–23.09) | $5.15 ($1.33–18.82) | $19.45 ($5.64–79.89) | $8.74 ($2.13–33.43) | $5.93 ($0.26–146.46) | $3.48 ($0.17–83.72) |
| Cameroon | LMI | AFR | $3.77 ($1.28–14.14) | $3.03 ($0.91–10.00) | $21.32 ($6.07–84.09) | $9.66 ($2.35–37.20) | $0.53 ($0.22–1.44) | $0.32 ($0.13–0.89) |
| Central African Republic | LI | AFR | $2.97 ($0.72–14.29) | $2.36 ($0.51–11.33) | $31.31 ($6.99–136.12) | $13.92 ($2.71–58.35) | $2.55 ($0.12–61.11) | $1.57 ($0.06–37.33) |
| Chad | LI | AFR | $2.87 ($0.92–11.17) | $2.35 ($0.71–9.02) | $28.11 ($7.33–116.18) | $12.64 ($2.74–49.74) | $1.96 ($0.09–44.47) | $1.15 ($0.05–26.31) |
| China | UMI | WPR | $3.98 ($1.37–14.23) | $3.21 ($1.12–9.94) | $17.83 ($5.02–74.04) | $7.93 ($1.89–30.47) | $5.31 ($0.21–149.03) | $3.19 ($0.14–87.55) |
| Colombia | UMI | AMR | $10.07 ($2.67–39.91) | $8.04 ($1.94–28.67) | $22.40 ($6.14–88.91) | $10.01 ($2.38–38.50) | $4.58 ($0.20–125.98) | $2.65 ($0.11–73.15) |
| Comoros | LMI | AFR | $9.96 ($1.86–49.02) | $7.95 ($1.26–43.04) | $14.32 ($3.93–63.75) | $6.43 ($1.56–27.07) | $3.94 ($0.18–97.83) | $2.33 ($0.10–56.59) |
| Congo | LMI | AFR | $7.68 ($1.95–31.37) | $6.24 ($1.38–24.94) | $26.75 ($7.04–108.31) | $12.08 ($2.59–47.40) | $4.30 ($0.21–107.87) | $2.57 ($0.11–73.64) |
| Congo DR | LI | AFR | $1.27 ($0.41–6.16) | $1.03 ($0.33–4.40) | $22.40 ($6.14–91.29) | $10.15 ($2.39–38.54) | $3.96 ($0.18–83.95) | $2.40 ($0.11–54.96) |
| Cote d'Ivoire | LMI | AFR | $4.83 ($1.64–17.67) | $3.89 ($1.17–12.53) | $19.21 ($5.48–76.54) | $8.78 ($2.16–34.27) | $2.68 ($0.14–57.53) | $1.64 ($0.09–33.14) |
| Cuba | UMI | AMR | $18.64 ($3.50–90.54) | $15.22 ($2.48–74.42) | $18.81 ($5.33–76.90) | $8.51 ($2.06–33.40) | $4.37 ($0.17–137.00) | $2.73 ($0.12–78.33) |
| Djibouti | LMI | EMR | $14.75 ($2.68–76.84) | $11.97 ($1.71–63.73) | $22.01 ($6.33–87.58) | $9.99 ($2.37–38.29) | $3.09 ($0.15–80.88) | $1.83 ($0.08–50.41) |
| Dominica | UMI | AMR | $24.09 ($3.81–135.02) | $19.54 ($2.42–119.16) | $19.75 ($5.57–79.78) | $8.80 ($2.11–32.91) | $3.20 ($0.15–91.39) | $2.00 ($0.08–61.15) |
| Dominican Republic | UMI | AMR | $15.15 ($3.29–66.34) | $12.14 ($2.30–54.17) | $16.35 ($4.46–69.83) | $7.33 ($1.73–28.24) | $3.01 ($0.12–84.91) | $1.93 ($0.08–52.94) |
| Ecuador | UMI | AMR | $12.36 ($3.12–50.75) | $9.97 ($2.05–40.41) | $20.59 ($5.73–83.05) | $9.24 ($2.22–34.72) | $3.87 ($0.16–108.64) | $2.39 ($0.09–69.33) |
| Egypt | LMI | EMR | $4.03 ($1.47–14.80) | $3.24 ($1.12–10.56) | $18.67 ($5.49–76.91) | $8.49 ($2.08–33.26) | $6.77 ($0.31–163.84) | $4.01 ($0.20–87.99) |
| El Salvador | UMI | AMR | $15.11 ($2.97–67.12) | $12.16 ($2.10–58.63) | $15.28 ($4.34–65.36) | $6.93 ($1.70–28.17) | $4.24 ($0.18–120.60) | $2.70 ($0.11–67.93) |
| Equatorial Guinea | UMI | AFR | $20.84 ($3.45–113.89) | $16.79 ($2.42–94.10) | $20.92 ($6.06–80.74) | $9.40 ($2.25–35.22) | $1.67 ($0.06–50.41) | $0.98 ($0.04–27.57) |
| Eritrea | LI | AFR | $4.40 ($0.98–22.21) | $3.60 ($0.67–18.29) | $24.55 ($6.61–96.57) | $11.08 ($2.61–41.80) | $7.94 ($0.33–196.86) | $5.03 ($0.22–114.52) |
| Ethiopia | LI | AFR | $16.06 ($2.87–83.73) | $12.66 ($1.95–68.54) | $20.52 ($5.91–83.47) | $9.25 ($2.12–34.89) | $3.46 ($0.16–93.06) | $2.06 ($0.09–52.65) |
| Fiji | UMI | WPR | $1.67 ($0.59–7.34) | $1.36 ($0.47–5.14) | $22.10 ($9.26–58.64) | $9.61 ($3.88–26.38) | $3.64 ($1.93–7.15) | $2.20 ($1.11–4.51) |
| Gabon | UMI | AFR | $21.17 ($3.40–115.57) | $16.89 ($2.33–98.61) | $21.82 ($5.99–85.11) | $9.84 ($2.30–37.19) | $4.20 ($0.17–121.05) | $2.61 ($0.11–71.94) |
| Gambia | LI | AFR | $21.31 ($3.59–113.60) | $17.15 ($2.64–89.89) | $30.21 ($7.09–129.31) | $13.69 ($2.91–56.48) | $2.95 ($0.11–106.03) | $1.72 ($0.07–58.30) |
| Georgia | UMI | EUR | $5.52 ($1.15–27.74) | $4.45 ($0.79–21.60) | $15.82 ($4.50–68.33) | $7.17 ($1.70–29.32) | $5.01 ($0.23–120.77) | $3.10 ($0.13–72.14) |
| Ghana | LMI | AFR | $23.42 ($3.69–128.80) | $18.78 ($2.48–110.41) | $20.93 ($5.75–84.48) | $9.35 ($2.21–35.38) | $4.25 ($0.16–122.54) | $2.44 ($0.10–72.44) |
| Grenada | UMI | AMR | $4.52 ($1.59–13.89) | $3.63 ($1.15–10.98) | $17.70 ($4.96–73.26) | $8.06 ($1.98–31.11) | $5.22 ($0.28–116.77) | $3.06 ($0.16–70.24) |
| Guatemala | UMI | AMR | $24.23 ($3.60–137.40) | $19.51 ($2.57–117.45) | $14.91 ($4.21–63.60) | $6.71 ($1.61–26.59) | $3.67 ($0.15–112.43) | $2.20 ($0.09–64.88) |
| Guinea | LMI | AFR | $10.57 ($2.79–40.46) | $8.35 ($1.95–31.83) | $17.31 ($4.89–69.42) | $7.74 ($1.87–32.33) | $4.33 ($0.19–120.35) | $2.61 ($0.11–70.88) |
| Guinea-Bissau | LI | AFR | $4.63 ($1.40–16.52) | $3.75 ($1.07–13.05) | $21.44 ($5.92–86.22) | $9.60 ($2.29–36.31) | $1.72 ($0.08–37.99) | $1.01 ($0.05–24.57) |
| Haiti | LMI | AMR | $6.22 ($1.32–30.66) | $4.99 ($0.88–26.09) | $20.51 ($5.72–83.33) | $9.10 ($2.24–34.84) | $3.19 ($0.14–76.34) | $1.99 ($0.09–49.33) |
| Honduras | LMI | AMR | $6.34 ($1.81–25.18) | $5.18 ($1.23–19.93) | $14.51 ($4.01–66.05) | $6.52 ($1.61–27.27) | $2.49 ($0.11–61.57) | $1.52 ($0.06–42.28) |
| India | LMI | SEAR | $8.58 ($2.68–26.75) | $6.89 ($1.90–22.90) | $19.41 ($5.44–79.47) | $8.74 ($2.10–32.55) | $5.11 ($0.21–132.90) | $3.12 ($0.14–90.18) |
| Indonesia | UMI | SEAR | $1.84 ($0.60–8.82) | $1.48 ($0.52–5.87) | $10.68 ($3.16–34.95) | $4.70 ($1.17–15.91) | $0.99 ($0.51–2.13) | $0.60 ($0.30–1.31) |
| Iran | UMI | EMR | $4.03 ($1.86–8.92) | $3.23 ($1.63–6.12) | $17.68 ($5.05–71.73) | $7.94 ($1.99–31.85) | $5.14 ($0.24–133.70) | $3.17 ($0.14–82.15) |
| Iraq | UMI | EMR | $6.17 ($1.95–22.33) | $5.03 ($1.50–16.77) | $21.58 ($6.02–85.90) | $9.71 ($2.27–36.51) | $5.92 ($0.29–146.84) | $3.56 ($0.17–81.25) |
| Jamaica | UMI | AMR | $7.49 ($2.82–23.86) | $6.03 ($2.14–17.64) | $19.03 ($5.36–79.26) | $8.61 ($2.13–32.95) | $4.83 ($0.22–134.46) | $2.91 ($0.14–68.31) |
| Jordan | LMI | EMR | $22.64 ($3.63–124.88) | $18.00 ($2.42–104.36) | $15.79 ($4.32–70.37) | $7.14 ($1.75–28.30) | $4.15 ($0.18–123.92) | $2.42 ($0.11–75.57) |
| Kazakhstan | UMI | EUR | $10.40 ($2.61–41.76) | $8.32 ($1.84–32.49) | $18.31 ($5.36–75.76) | $8.27 ($2.08–32.42) | $5.19 ($0.24–141.09) | $3.07 ($0.14–89.83) |
| Kenya | LMI | AFR | $11.97 ($2.98–46.45) | $9.57 ($2.17–36.66) | $31.77 ($7.36–140.13) | $14.29 ($2.77–60.49) | $4.74 ($0.20–130.51) | $2.93 ($0.11–78.27) |
| Kiribati | LMI | WPR | $4.00 ($1.81–11.71) | $3.22 ($1.37–9.06) | $19.38 ($5.37–79.67) | $8.77 ($2.15–32.85) | $5.59 ($2.89–12.40) | $3.38 ($1.66–7.87) |
| Korea DPR | LI | SEAR | $11.47 ($2.12–57.97) | $9.19 ($1.41–50.10) | $17.48 ($4.94–71.18) | $7.76 ($1.98–31.89) | $3.64 ($0.16–83.06) | $2.07 ($0.10–48.90) |
| Kyrgyz Republic | LMI | EUR | $4.51 ($1.37–16.99) | $3.65 ($0.95–13.66) | $16.37 ($4.73–68.89) | $7.38 ($1.83–30.10) | $7.43 ($0.31–169.96) | $4.77 ($0.22–119.11) |
| Lao PDR | LMI | WPR | $7.93 ($1.96–32.83) | $6.33 ($1.40–26.38) | $23.47 ($6.31–95.19) | $10.59 ($2.43–40.97) | $6.42 ($0.27–167.38) | $3.84 ($0.17–96.05) |
| Lebanon | LMI | EMR | $7.25 ($1.87–29.00) | $5.85 ($1.31–24.26) | $21.30 ($6.44–64.95) | $9.50 ($2.31–32.57) | $4.39 ($0.19–110.62) | $2.55 ($0.12–70.60) |
| Lesotho | LMI | AFR | $11.69 ($2.52–50.91) | $9.34 ($1.69–45.18) | $13.97 ($3.81–63.35) | $6.22 ($1.55–25.34) | $3.34 ($0.12–100.25) | $1.97 ($0.07–70.48) |
| Liberia | LI | AFR | $6.34 ($1.29–32.36) | $5.14 ($0.87–27.68) | $20.18 ($5.78–83.87) | $9.02 ($2.16–33.67) | $4.34 ($0.21–95.49) | $2.58 ($0.12–65.97) |
| Libya | UMI | EMR | $4.24 ($1.06–18.72) | $3.38 ($0.75–14.88) | $21.64 ($6.07–87.00) | $9.63 ($2.31–36.02) | $3.59 ($0.16–82.76) | $2.13 ($0.09–51.66) |
| North Macedonia | UMI | EUR | $15.07 ($3.08–66.08) | $12.06 ($2.28–54.17) | $35.61 ($7.66–156.86) | $15.94 ($2.84–75.30) | $3.94 ($0.15–115.88) | $2.38 ($0.10–66.11) |
| Madagascar | LI | AFR | $1.83 ($0.57–8.27) | $1.48 ($0.41–6.12) | $21.76 ($5.95–84.97) | $9.79 ($2.27–37.00) | $4.62 ($0.20–99.90) | $2.64 ($0.13–58.35) |
| Malawi | LI | AFR | $1.97 ($0.60–9.29) | $1.61 ($0.42–6.97) | $16.50 ($4.74–69.74) | $7.46 ($1.92–29.39) | $4.20 ($2.27–7.96) | $2.53 ($1.32–5.03) |
| Malaysia | UMI | WPR | $11.56 ($2.96–47.48) | $9.37 ($2.20–36.55) | $19.16 ($5.52–78.62) | $8.52 ($2.03–32.88) | $4.47 ($0.20–106.19) | $2.65 ($0.12–71.92) |
| Maldives | UMI | SEAR | $24.35 ($3.91–139.30) | $19.50 ($2.54–115.58) | $13.79 ($3.70–63.79) | $6.23 ($1.49–25.96) | $2.28 ($0.05–99.76) | $1.40 ($0.04–65.97) |
| Mali | LI | AFR | $2.85 ($0.97–11.86) | $2.31 ($0.71–8.50) | $26.83 ($6.72–113.69) | $11.82 ($2.52–45.67) | $2.60 ($0.13–54.54) | $1.55 ($0.07–33.37) |
| Marshall Islands | UMI | WPR | $23.36 ($3.49–128.33) | $18.64 ($2.49–108.73) | $16.51 ($4.75–69.31) | $7.40 ($1.81–28.97) | $3.46 ($0.15–100.92) | $2.03 ($0.08–57.82) |
| Mauritania | LMI | AFR | $7.22 ($1.89–29.12) | $5.81 ($1.24–24.00) | $35.03 ($7.50–156.31) | $15.54 ($2.71–71.89) | $4.99 ($0.22–124.68) | $3.00 ($0.14–75.96) |
| Mauritius | UMI | AFR | $24.35 ($3.70–138.60) | $19.49 ($2.47–116.03) | $14.02 ($3.81–64.09) | $6.26 ($1.52–26.83) | $3.18 ($0.14–91.83) | $1.92 ($0.08–60.16) |
| Mexico | UMI | AMR | $7.10 ($2.15–26.96) | $5.78 ($1.65–20.75) | $20.75 ($5.53–82.01) | $9.42 ($2.19–35.21) | $1.45 ($0.42–4.87) | $0.88 ($0.24–3.04) |
| Micronesia | LMI | WPR | $16.54 ($2.80–86.15) | $13.23 ($1.94–74.97) | $17.29 ($4.85–72.73) | $7.79 ($1.99–31.25) | $4.47 ($0.20–120.36) | $2.71 ($0.11–71.51) |
| Moldova | UMI | EUR | $25.41 ($6.19–114.11) | $20.36 ($4.21–97.43) | $19.73 ($5.71–79.05) | $8.92 ($2.18–32.65) | $4.16 ($0.18–126.12) | $2.55 ($0.10–73.19) |
| Mongolia | UMI | WPR | $18.81 ($3.31–90.76) | $14.95 ($2.27–78.60) | $35.98 ($7.41–156.70) | $15.87 ($2.94–73.21) | $4.89 ($0.19–144.62) | $2.83 ($0.11–80.34) |
| Montenegro | UMI | EUR | $24.37 ($3.64–139.39) | $19.64 ($2.51–119.55) | $22.56 ($6.20–88.72) | $10.02 ($2.39–37.25) | $4.01 ($0.15–128.23) | $2.43 ($0.09–74.53) |
| Morocco | LMI | EMR | $6.84 ($2.17–25.37) | $5.54 ($1.49–18.39) | $19.81 ($5.77–76.85) | $8.90 ($2.13–33.77) | $5.92 ($0.26–145.15) | $3.46 ($0.17–82.92) |
| Mozambique | LI | AFR | $2.14 ($0.73–11.80) | $1.75 ($0.57–8.71) | $22.73 ($6.34–88.27) | $10.28 ($2.39–38.98) | $4.45 ($2.81–7.35) | $2.69 ($1.62–4.67) |
| Myanmar | LMI | SEAR | $3.19 ($1.09–12.51) | $2.59 ($0.82–9.03) | $20.01 ($5.57–79.70) | $9.04 ($2.21–34.59) | $5.38 ($0.28–126.36) | $3.14 ($0.16–65.11) |
| Namibia | LMI | AFR | $14.03 ($2.85–62.55) | $11.14 ($1.94–52.72) | $35.69 ($7.57–151.73) | $15.75 ($2.95–74.26) | $3.46 ($0.15–95.47) | $2.12 ($0.09–56.52) |
| Nepal | LMI | SEAR | $3.88 ($1.23–14.67) | $3.11 ($0.91–11.03) | $16.77 ($4.77–71.41) | $7.46 ($1.89–29.05) | $1.21 ($0.62–2.62) | $0.73 ($0.37–1.63) |
| Nicaragua | LMI | AMR | $9.13 ($2.26–37.45) | $7.46 ($1.53–31.02) | $21.56 ($6.07–85.85) | $9.56 ($2.21–38.38) | $6.07 ($0.27–148.55) | $3.69 ($0.16–104.04) |
| Niger | LI | AFR | $2.23 ($0.98–6.27) | $1.79 ($0.73–4.91) | $16.40 ($4.48–50.29) | $7.11 ($1.65–26.56) | $1.99 ($0.08–48.07) | $1.20 ($0.05–29.76) |
| Nigeria | LMI | AFR | $1.23 ($0.41–5.80) | $0.98 ($0.34–4.10) | $16.26 ($4.63–69.60) | $7.33 ($1.84–28.55) | $0.10 ($0.04–0.22) | $0.06 ($0.03–0.14) |
| Pakistan | LMI | EMR | $24.43 ($3.80–139.04) | $19.60 ($2.63–118.44) | $20.50 ($5.66–82.39) | $9.26 ($2.32–35.72) | $4.25 ($0.17–128.95) | $2.59 ($0.10–76.88) |
| Papua New Guinea | LMI | WPR | $1.78 ($0.64–7.66) | $1.45 ($0.54–5.26) | $15.25 ($4.39–66.93) | $6.91 ($1.70–26.14) | $4.12 ($0.20–96.90) | $2.38 ($0.10–53.14) |
| Paraguay | UMI | AMR | $7.89 ($2.17–30.09) | $6.41 ($1.52–24.18) | $25.85 ($6.55–104.20) | $11.50 ($2.50–45.79) | $3.38 ($0.13–88.31) | $1.97 ($0.08–66.02) |
| Peru | UMI | AMR | $15.05 ($3.13–66.02) | $12.07 ($2.29–55.49) | $27.21 ($7.04–112.99) | $12.07 ($2.62–48.80) | $4.36 ($0.19–137.46) | $2.68 ($0.11–85.34) |
| Philippines | LMI | WPR | $10.98 ($2.73–43.48) | $8.95 ($1.97–34.24) | $20.82 ($5.69–70.56) | $9.29 ($2.09–33.40) | $4.21 ($0.19–119.75) | $2.48 ($0.11–65.76) |
| Rwanda | LI | AFR | $4.81 ($1.72–17.31) | $3.91 ($1.28–12.47) | $18.81 ($6.67–64.70) | $8.27 ($3.14–26.05) | $2.62 ($1.78–3.96) | $1.58 ($1.03–2.54) |
| Samoa | UMI | WPR | $3.50 ($1.08–14.68) | $2.82 ($0.75–10.95) | $10.18 ($3.99–27.43) | $4.45 ($1.50–13.68) | $5.28 ($0.25–114.04) | $3.17 ($0.14–74.20) |
| Sao Tome and Principe | LMI | AFR | $18.09 ($3.18–95.94) | $14.66 ($2.18–81.54) | $20.28 ($5.67–81.75) | $9.12 ($2.14–34.90) | $4.71 ($0.21–138.87) | $2.85 ($0.13–87.39) |
| Senegal | LMI | AFR | $14.35 ($2.49–73.17) | $11.47 ($1.66–63.35) | $16.18 ($4.70–69.72) | $7.26 ($1.76–28.23) | $5.28 ($0.23–138.70) | $3.13 ($0.14–80.09) |
| Serbia | UMI | EUR | $4.51 ($1.41–17.75) | $3.63 ($1.03–12.60) | $23.40 ($9.85–61.81) | $10.20 ($4.14–28.45) | $5.54 ($0.27–130.12) | $3.32 ($0.16–70.86) |
| Sierra Leone | LI | AFR | $21.76 ($3.70–117.18) | $17.47 ($2.55–94.87) | $20.20 ($5.60–77.83) | $9.09 ($2.15–33.53) | $4.28 ($0.18–137.64) | $2.59 ($0.11–79.07) |
| Solomon Islands | LMI | WPR | $3.76 ($1.03–16.63) | $3.07 ($0.74–12.75) | $18.54 ($5.19–78.78) | $8.43 ($2.06–32.93) | $1.07 ($0.79–1.46) | $0.64 ($0.45–0.93) |
| Somalia | LI | EMR | $11.05 ($2.05–57.61) | $8.81 ($1.43–47.92) | $24.93 ($6.72–99.71) | $11.10 ($2.57–42.13) | $6.79 ($0.29–189.75) | $3.96 ($0.18–102.70) |
| South Africa | UMI | AFR | $2.21 ($0.69–9.92) | $1.80 ($0.49–7.24) | $24.59 ($6.71–94.93) | $11.18 ($2.52–40.38) | $2.18 ($0.10–52.30) | $1.32 ($0.06–30.93) |
| South Sudan | LI | AFR | $7.40 ($2.26–26.76) | $6.00 ($1.65–20.69) | $21.94 ($6.06–85.51) | $9.82 ($2.38–36.88) | $3.61 ($0.16–90.57) | $2.24 ($0.11–58.62) |
| Sri Lanka | LMI | SEAR | $3.94 ($1.11–16.31) | $3.19 ($0.80–12.57) | $27.08 ($6.90–109.82) | $12.14 ($2.54–49.51) | $1.97 ($0.09–47.71) | $1.22 ($0.05–28.61) |
| St. Lucia | UMI | AMR | $9.10 ($2.45–35.92) | $7.32 ($1.70–27.55) | $11.08 ($5.07–25.82) | $4.89 ($1.81–13.76) | $4.79 ($0.23–109.07) | $2.77 ($0.13–74.67) |
| St. Vincent and the Grenadines | UMI | AMR | $24.54 ($3.72–135.79) | $19.52 ($2.59–115.67) | $15.51 ($4.27–66.94) | $6.95 ($1.73–27.53) | $3.97 ($0.17–116.69) | $2.36 ($0.10–71.05) |
| Sudan | LI | EMR | $24.26 ($3.70–136.61) | $19.50 ($2.58–119.32) | $15.97 ($4.64–67.38) | $7.16 ($1.80–27.90) | $4.14 ($0.16–116.20) | $2.52 ($0.11–68.97) |
| Suriname | UMI | AMR | $2.19 ($0.78–8.91) | $1.79 ($0.59–6.65) | $25.01 ($6.57–103.34) | $11.15 ($2.39–42.42) | $4.73 ($0.24–96.04) | $2.77 ($0.14–63.74) |
| Eswatini | LMI | AFR | $23.29 ($3.53–127.28) | $18.57 ($2.44–112.37) | $35.93 ($7.41–167.14) | $15.99 ($2.83–69.97) | $3.76 ($0.15–123.41) | $2.30 ($0.09–71.35) |
| Syrian Arab Republic | LI | EMR | $2.98 ($0.89–12.58) | $2.41 ($0.64–9.68) | $18.45 ($5.28–76.42) | $8.31 ($2.00–31.22) | $8.18 ($0.37–189.54) | $5.00 ($0.22–116.40) |
| Tajikistan | LMI | EUR | $4.68 ($1.35–19.05) | $3.80 ($0.90–15.11) | $20.51 ($5.79–81.40) | $9.22 ($2.18–34.04) | $7.42 ($0.32–175.32) | $4.44 ($0.20–112.18) |
| Tanzania | LMI | AFR | $1.84 ($0.88–3.77) | $1.46 ($0.66–3.23) | $33.94 ($11.01–126.10) | $15.05 ($4.88–52.64) | $6.57 ($0.32–142.13) | $3.92 ($0.20–86.42) |
| Thailand | UMI | SEAR | $10.07 ($2.71–39.03) | $8.08 ($1.95–29.05) | $18.06 ($5.32–74.05) | $8.11 ($1.91–30.69) | $4.33 ($0.20–118.50) | $2.62 ($0.12–69.81) |
| Timor-Leste | LMI | SEAR | $8.36 ($1.56–42.78) | $6.71 ($1.12–36.53) | $19.17 ($5.51–76.72) | $8.82 ($2.11–33.34) | $4.53 ($0.20–105.16) | $2.59 ($0.12–60.34) |
| Togo | LI | AFR | $4.05 ($1.13–16.77) | $3.23 ($0.77–12.70) | $17.30 ($4.90–71.24) | $7.81 ($1.94–29.43) | $0.07 ($0.03–0.18) | $0.04 ($0.02–0.12) |
| Tonga | UMI | WPR | $18.16 ($3.10–95.58) | $14.44 ($2.16–82.88) | $17.95 ($5.17–69.87) | $8.04 ($1.94–30.01) | $5.09 ($0.23–129.27) | $3.12 ($0.13–82.59) |
| Tunisia | LMI | EMR | $10.81 ($2.49–44.59) | $8.64 ($1.82–34.91) | $20.23 ($5.87–79.12) | $9.16 ($2.15–34.74) | $5.57 ($0.22–153.49) | $3.38 ($0.14–90.63) |
| Turkiye | UMI | EUR | $8.67 ($2.49–33.32) | $7.03 ($1.93–24.03) | $18.71 ($5.15–76.04) | $8.47 ($2.10–32.09) | $4.79 ($0.22–124.30) | $2.86 ($0.13–70.06) |
| Turkmenistan | UMI | EUR | $16.15 ($3.38–71.73) | $12.94 ($2.22–58.31) | $27.76 ($6.94–113.42) | $12.35 ($2.57–49.18) | $3.35 ($0.14–93.89) | $2.00 ($0.08–59.69) |
| Tuvalu | UMI | WPR | $20.99 ($3.41–120.20) | $16.88 ($2.35–96.74) | $15.27 ($4.41–68.24) | $6.89 ($1.73–28.59) | $4.17 ($0.19–123.12) | $2.45 ($0.10–72.56) |
| Uganda | LI | AFR | $1.98 ($0.72–5.41) | $1.60 ($0.56–4.22) | $16.65 ($5.40–58.37) | $7.46 ($2.15–26.25) | $6.49 ($0.35–142.19) | $3.82 ($0.18–78.28) |
| Ukraine | UMI | EUR | $10.89 ($2.67–42.54) | $8.93 ($1.85–35.48) | $20.49 ($5.66–85.69) | $9.22 ($2.24–35.72) | $4.79 ($0.20–134.21) | $2.90 ($0.13–84.16) |
| Uzbekistan | LMI | EUR | $5.41 ($1.76–18.87) | $4.33 ($1.33–14.43) | $20.13 ($5.54–79.24) | $9.05 ($2.14–33.44) | $6.25 ($0.31–143.13) | $3.83 ($0.18–81.95) |
| Vanuatu | LMI | WPR | $14.89 ($2.70–75.74) | $11.89 ($1.75–67.69) | $25.34 ($6.56–98.82) | $11.17 ($2.57–43.35) | $4.94 ($0.21–139.34) | $3.08 ($0.13–93.93) |
| Venezuela | UMI | AMR | $11.94 ($2.86–49.42) | $9.56 ($2.12–35.80) | $24.25 ($6.65–98.69) | $10.90 ($2.50–41.13) | $3.45 ($0.13–116.14) | $2.16 ($0.09–69.29) |
| Viet Nam | LMI | WPR | $5.83 ($1.97–20.92) | $4.71 ($1.47–15.62) | $15.38 ($4.31–64.00) | $6.88 ($1.74–27.57) | $5.11 ($0.26–118.98) | $3.17 ($0.16–75.63) |
| Yemen | LI | EMR | $1.42 ($0.43–6.94) | $1.16 ($0.33–5.41) | $20.50 ($5.81–82.04) | $9.22 ($2.17–34.64) | $7.30 ($0.31–159.93) | $4.42 ($0.21–104.80) |
| Zambia | LMI | AFR | $3.13 ($0.99–10.07) | $2.53 ($0.68–7.83) | $24.99 ($6.69–98.02) | $11.27 ($2.53–43.50) | $0.97 ($0.43–2.56) | $0.58 ($0.26–1.56) |
| Zimbabwe | LMI | AFR | $5.92 ($1.80–22.08) | $4.81 ($1.29–16.74) | $23.16 ($5.91–88.85) | $10.16 ($2.33–39.47) | $4.26 ($0.21–110.42) | $2.70 ($0.13–58.10) |

^a^ LI: Gross national income (GNI) per capita of $1,135 or less in 2024; LMI: GNI per capita $1,136 and $4,495; UMI: GNI per capita of $4,496 and $13,935 (World Bank 2024) [18].

Note: AFR = African region; AMR = Region of the Americas; DTP3 = diphtheria-tetanus-pertussis third dose coverage; EMR = Eastern Mediterranean region; EUR = European region; GDP = gross domestic product; LI = low-income; LMI = lower middle-income; SEAR = Southeast Asian region; UMI = upper middle-income; WPR = Western Pacific region.

**Appendix Table E. Predicted incremental cost per dose in 2024 by delivery modality and country.**

| **Country** | **Income** | **Region** | **Economic Cost of Childhood Vaccine Delivery** | **Financial Cots of Childhood Vaccine Delivery** | **Economic Cost of Adolescent Vaccine Delivery** | **Financial Cost of Adolescent Vaccine Delivery** | **Economic Cost of Campaign Vaccine Delivery** | **Financial Cost of Campaign Vaccine Delivery** |
| --- | --- | --- | --- | --- | --- | --- | --- | --- |
| Afghanistan | LI | EMR | $5.11 ($1.21–19.19) | $2.79 ($0.67–10.36) | $25.52 ($8.59–65.75) | $18.12 ($6.29–46.07) | $3.15 ($0.90–14.39) | $2.14 ($0.63–9.13) |
| Albania | UMI | EUR | $33.70 ($12.41–90.48) | $18.43 ($7.70–44.30) | $16.29 ($6.47–37.71) | $11.53 ($4.57–26.84) | $7.47 ($2.83–19.70) | $5.09 ($1.89–13.60) |
| Algeria | UMI | AFR | $26.11 ($11.78–59.71) | $14.28 ($7.12–30.16) | $19.38 ($7.54–45.25) | $13.81 ($5.30–32.32) | $6.83 ($3.02–16.65) | $4.68 ($2.12–10.81) |
| Angola | LMI | AFR | $10.72 ($3.60–30.69) | $5.88 ($2.07–16.29) | $24.63 ($8.42–62.85) | $17.53 ($6.13–44.35) | $3.40 ($1.03–14.51) | $2.31 ($0.73–9.31) |
| Argentina | UMI | AMR | $41.79 ($8.07–181.79) | $22.75 ($4.71–98.11) | $31.72 ($8.63–102.09) | $22.48 ($6.16–71.57) | $5.28 ($1.87–16.61) | $3.60 ($1.31–11.06) |
| Armenia | UMI | EUR | $35.27 ($12.65–95.55) | $19.14 ($7.82–47.69) | $18.19 ($7.06–42.05) | $12.91 ($5.04–30.17) | $7.27 ($2.79–19.21) | $4.97 ($1.88–13.16) |
| Azerbaijan | UMI | EUR | $43.98 ($7.97–208.32) | $24.08 ($4.71–108.65) | $31.72 ($8.63–102.09) | $22.48 ($6.16–71.57) | $3.73 ($0.90–19.20) | $2.56 ($0.61–12.51) |
| Bangladesh | LMI | SEAR | $14.92 ($7.49–33.55) | $8.19 ($4.34–17.35) | $14.98 ($6.30–33.20) | $10.66 ($4.55–23.49) | $0.54 ($0.14–2.36) | $0.37 ($0.10–1.58) |
| Belarus | UMI | EUR | $30.83 ($12.39–77.28) | $16.82 ($7.71–38.12) | $15.66 ($6.24–36.42) | $11.12 ($4.40–26.06) | $8.37 ($3.11–22.27) | $5.72 ($2.09–15.32) |
| Belize | UMI | AMR | $42.28 ($11.45–146.07) | $23.17 ($6.93–74.44) | $22.70 ($8.11–57.12) | $16.16 ($5.79–39.99) | $7.70 ($2.98–20.20) | $5.26 ($2.02–13.72) |
| Benin | LMI | AFR | $6.69 ($2.23–19.54) | $3.64 ($1.28–10.29) | $23.52 ($8.20–60.15) | $16.78 ($5.93–42.38) | $3.09 ($0.88–13.49) | $2.11 ($0.59–8.76) |
| Bhutan | LMI | SEAR | $29.92 ($10.22–85.74) | $16.34 ($6.38–42.47) | $15.38 ($6.25–35.54) | $10.94 ($4.43–25.12) | $8.81 ($3.23–23.46) | $6.02 ($2.18–16.15) |
| Bolivia | LMI | AMR | $38.50 ($7.75–165.78) | $21.05 ($4.47–88.09) | $31.12 ($8.66–97.27) | $22.08 ($6.22–68.39) | $4.65 ($1.14–23.15) | $3.18 ($0.78–15.27) |
| Bosnia and Herzegovina | UMI | EUR | $47.21 ($9.04–215.64) | $25.97 ($5.33–112.33) | $31.72 ($8.63–102.09) | $22.48 ($6.16–71.57) | $5.85 ($2.12–17.71) | $4.02 ($1.44–11.70) |
| Botswana | UMI | AFR | $21.14 ($8.21–54.02) | $11.58 ($5.03–26.83) | $15.17 ($6.57–33.04) | $10.84 ($4.70–23.28) | $8.80 ($3.32–23.00) | $6.02 ($2.23–15.83) |
| Brazil | UMI | AMR | $27.43 ($12.21–63.74) | $15.05 ($7.19–33.10) | $20.22 ($7.63–48.53) | $14.46 ($5.41–34.75) | $5.70 ($2.43–14.87) | $3.91 ($1.79–9.53) |
| Burkina Faso | LI | AFR | $3.34 ($1.40–7.82) | $1.83 ($0.80–4.08) | $15.19 ($6.24–35.82) | $10.82 ($4.51–25.19) | $2.95 ($1.26–8.40) | $2.01 ($0.82–6.33) |
| Burundi | LI | AFR | $4.77 ($1.85–12.47) | $2.62 ($1.07–6.32) | $18.12 ($7.77–39.71) | $12.86 ($5.57–28.06) | $2.50 ($0.92–7.39) | $1.71 ($0.62–5.06) |
| Cabo Verde | UMI | AFR | $38.69 ($12.20–117.43) | $20.99 ($7.54–58.72) | $18.84 ($7.26–44.13) | $13.40 ($5.17–31.48) | $6.76 ($2.66–17.63) | $4.63 ($1.79–12.25) |
| Cambodia | LMI | WPR | $26.67 ($8.24–78.28) | $14.59 ($4.84–41.16) | $32.92 ($15.27–76.65) | $23.40  ($10.96–54.85) | $5.59 ($2.29–15.06) | $3.84 ($1.56–9.85) |
| Cameroon | LMI | AFR | $8.69 ($3.21–23.01) | $4.75 ($1.86–12.12) | $24.33 ($8.39–62.12) | $17.34 ($6.10–43.70) | $4.36 ($1.88–11.34) | $2.98 ($1.30–7.55) |
| Central African Republic | LI | AFR | $2.80 ($0.85–8.82) | $1.54 ($0.49–4.65) | $23.31 ($8.06–59.66) | $16.60 ($5.85–42.18) | $4.05 ($1.13–17.54) | $2.76 ($0.76–11.55) |
| Chad | LI | AFR | $3.11 ($1.21–8.32) | $1.70 ($0.67–4.48) | $23.31 ($8.06–59.66) | $16.60 ($5.85–42.18) | $3.75 ($1.19–14.45) | $2.57 ($0.81–9.50) |
| China | UMI | WPR | $22.06 ($10.52–53.44) | $12.05 ($5.89–28.43) | $16.29 ($6.47–37.71) | $11.53 ($4.57–26.84) | $3.87 ($1.42–12.39) | $2.64 ($1.08–8.02) |
| Colombia | UMI | AMR | $31.36 ($12.32–78.43) | $17.06 ($7.30–39.81) | $21.84 ($7.95–53.87) | $15.60 ($5.66–38.02) | $6.36 ($2.84–15.16) | $4.35 ($1.97–10.09) |
| Comoros | LMI | AFR | $20.16 ($3.76–94.62) | $11.07 ($2.21–49.16) | $27.55 ($8.84–74.30) | $19.56 ($6.43–52.85) | $2.43 ($0.73–9.63) | $1.66 ($0.49–6.54) |
| Congo | LMI | AFR | $20.83 ($5.73–68.85) | $11.35 ($3.35–36.12) | $27.40 ($8.83–73.79) | $19.46 ($6.43–52.42) | $6.20 ($2.30–18.28) | $4.26 ($1.55–12.14) |
| Congo DR | LI | AFR | $3.52 ($1.12–10.25) | $1.92 ($0.63–5.61) | $23.89 ($8.28–60.85) | $16.99 ($6.03–42.97) | $2.78 ($0.77–12.80) | $1.90 ($0.57–8.23) |
| Cote d'Ivoire | LMI | AFR | $11.90 ($4.36–32.47) | $6.50 ($2.49–17.09) | $24.34 ($8.40–62.13) | $17.34 ($6.10–43.73) | $4.01 ($1.74–10.63) | $2.75 ($1.19–6.99) |
| Cuba | UMI | AMR | $30.38 ($12.47–74.94) | $16.57 ($7.72–37.26) | $15.66 ($6.24–36.42) | $11.12 ($4.40–26.06) | $7.39 ($2.81–19.47) | $5.04 ($1.88–13.44) |
| Djibouti | LMI | EMR | $25.31 ($5.97–97.20) | $13.87 ($3.61–50.69) | $26.10 ($8.73–67.88) | $18.54 ($6.24–47.68) | $5.54 ($2.14–15.80) | $3.82 ($1.44–10.63) |
| Dominica | UMI | AMR | $28.17 ($9.05–83.81) | $15.32 ($5.47–41.53) | $18.78 ($7.85–41.13) | $13.32 ($5.58–29.00) | $6.91 ($2.76–17.81) | $4.71 ($1.85–12.03) |
| Dominican Republic | UMI | AMR | $26.49 ($9.97–68.34) | $14.46 ($5.99–34.05) | $21.56 ($8.40–50.49) | $15.31 ($5.94–35.60) | $4.84 ($1.90–13.23) | $3.33 ($1.29–9.02) |
| Ecuador | UMI | AMR | $43.59 ($8.14–198.77) | $23.73 ($4.73–106.27) | $31.72 ($8.63–102.09) | $22.48 ($6.16–71.57) | $4.71 ($1.41–18.37) | $3.22 ($0.98–12.07) |
| Egypt | LMI | EMR | $23.72 ($11.60–54.40) | $13.01 ($6.84–28.03) | $16.29 ($6.47–37.71) | $11.53 ($4.57–26.84) | $5.27 ($2.35–13.12) | $3.60 ($1.69–8.49) |
| El Salvador | UMI | AMR | $32.23 ($12.84–80.77) | $17.53 ($7.92–40.00) | $17.52 ($6.84–40.32) | $12.41 ($4.87–28.93) | $4.94 ($1.87–13.60) | $3.37 ($1.25–9.37) |
| Equatorial Guinea | UMI | AFR | $16.59 ($4.45–59.30) | $9.09 ($2.59–30.60) | $24.07 ($8.34–61.21) | $17.15 ($6.08–43.23) | $4.27 ($1.40–15.52) | $2.91 ($0.93–10.14) |
| Eritrea | LI | AFR | $6.85 ($2.53–19.08) | $3.77 ($1.53–9.57) | $15.59 ($6.74–34.04) | $11.10 ($4.82–23.96) | $8.64 ($3.22–22.89) | $5.91 ($2.16–15.77) |
| Ethiopia | LI | AFR | $24.58 ($7.64–74.95) | $13.37 ($4.58–37.66) | $22.22 ($8.47–52.22) | $15.72 ($6.13–36.29) | $6.19 ($2.55–15.74) | $4.24 ($1.72–10.94) |
| Fiji | UMI | WPR | $8.28 ($2.16–28.53) | $4.55 ($1.21–15.54) | $26.52 ($8.79–69.85) | $18.86 ($6.34–49.27) | $2.77 ($0.96–9.20) | $1.88 ($0.70–5.96) |
| Gabon | UMI | AFR | $28.87 ($10.02–80.19) | $15.77 ($6.17–40.02) | $16.95 ($7.14–37.16) | $12.03 ($5.05–26.04) | $8.11 ($3.05–21.41) | $5.55 ($2.07–14.74) |
| Gambia | LI | AFR | $34.57 ($6.92–150.31) | $18.92 ($4.16–78.75) | $28.73 ($8.85–80.64) | $20.32 ($6.43–57.30) | $4.65 ($1.14–23.17) | $3.19 ($0.78–15.28) |
| Georgia | UMI | EUR | $9.57 ($2.32–35.33) | $5.22 ($1.35–18.36) | $25.93 ($8.77–66.95) | $18.39 ($6.32–47.30) | $3.90 ($1.45–11.52) | $2.66 ($0.99–7.90) |
| Ghana | LMI | AFR | $39.77 ($11.27–129.81) | $21.79 ($6.80–66.55) | $24.50 ($8.32–64.45) | $17.40 ($5.91–45.42) | $6.71 ($2.63–17.86) | $4.58 ($1.79–12.11) |
| Grenada | UMI | AMR | $12.44 ($6.18–25.43) | $6.85 ($3.55–13.10) | $15.41 ($6.67–33.64) | $10.99 ($4.77–23.73) | $5.67 ($2.61–13.26) | $3.88 ($1.79–8.79) |
| Guatemala | UMI | AMR | $37.37 ($11.86–112.63) | $20.28 ($7.35–56.54) | $18.84 ($7.26–44.13) | $13.40 ($5.17–31.48) | $4.37 ($1.67–12.01) | $2.98 ($1.10–8.44) |
| Guinea | LMI | AFR | $34.63 ($9.57–113.07) | $18.96 ($5.69–58.92) | $29.45 ($8.50–88.77) | $20.90 ($6.15–63.00) | $4.63 ($1.84–13.05) | $3.18 ($1.28–8.63) |
| Guinea-Bissau | LI | AFR | $5.33 ($1.94–14.88) | $2.91 ($1.10–7.91) | $23.31 ($8.06–59.66) | $16.60 ($5.85–42.18) | $3.39 ($0.95–14.83) | $2.32 ($0.64–9.85) |
| Haiti | LMI | AMR | $6.71 ($1.68–24.24) | $3.69 ($0.98–12.73) | $24.16 ($8.36–61.66) | $17.21 ($6.10–43.22) | $4.20 ($1.37–15.29) | $2.87 ($0.92–10.01) |
| Honduras | LMI | AMR | $14.48 ($4.00–47.62) | $7.88 ($2.33–25.13) | $25.55 ($8.60–65.90) | $18.15 ($6.29–46.16) | $1.14 ($0.35–4.10) | $0.78 ($0.22–3.10) |
| India | LMI | SEAR | $44.05 ($8.08–205.57) | $24.03 ($4.76–108.89) | $31.72 ($8.63–102.09) | $22.48 ($6.16–71.57) | $5.07 ($1.71–17.04) | $3.48 ($1.16–11.37) |
| Indonesia | UMI | SEAR | $14.43 ($7.60–29.65) | $7.94 ($4.21–15.95) | $17.16 ($7.12–37.96) | $12.16 ($5.06–26.71) | $0.97 ($0.48–2.17) | $0.66 ($0.32–1.51) |
| Iran | UMI | EMR | $29.91 ($6.87–107.00) | $16.47 ($3.94–57.88) | $31.72 ($8.63–102.09) | $22.48 ($6.16–71.57) | $3.19 ($1.14–10.58) | $2.17 ($0.86–6.74) |
| Iraq | UMI | EMR | $25.40 ($12.05–58.82) | $13.92 ($7.22–29.96) | $15.66 ($6.24–36.42) | $11.12 ($4.40–26.06) | $6.52 ($2.88–15.86) | $4.45 ($1.99–10.35) |
| Jamaica | UMI | AMR | $26.45 ($11.54–61.36) | $14.45 ($6.84–31.47) | $20.79 ($7.80–49.82) | $14.84 ($5.55–35.19) | $5.43 ($2.51–12.87) | $3.72 ($1.79–8.35) |
| Jordan | LMI | EMR | $31.11 ($11.64–82.31) | $16.96 ($7.24–40.21) | $15.66 ($6.24–36.42) | $11.12 ($4.40–26.06) | $5.37 ($2.06–14.54) | $3.66 ($1.37–10.03) |
| Kazakhstan | UMI | EUR | $29.74 ($12.84–70.37) | $16.18 ($7.88–35.44) | $16.93 ($6.68–38.86) | $11.95 ($4.73–27.86) | $7.08 ($2.71–18.66) | $4.83 ($1.80–12.89) |
| Kenya | LMI | AFR | $27.44 ($12.43–63.79) | $14.95 ($7.58–32.48) | $15.66 ($6.24–36.42) | $11.12 ($4.40–26.06) | $8.26 ($3.32–20.79) | $5.64 ($2.21–14.13) |
| Kiribati | LMI | WPR | $12.49 ($6.17–26.18) | $6.82 ($3.61–13.55) | $17.59 ($7.54–38.05) | $12.53 ($5.39–26.94) | $5.52 ($2.27–15.69) | $3.77 ($1.68–10.02) |
| Korea DPR | LI | SEAR | $13.17 ($4.53–36.81) | $7.19 ($2.81–18.36) | $14.64 ($6.27–32.92) | $10.44 ($4.49–23.14) | $6.13 ($2.46–15.60) | $4.20 ($1.60–10.79) |
| Kyrgyz Republic | LMI | EUR | $14.34 ($6.52–33.37) | $7.90 ($4.02–16.55) | $15.66 ($6.24–36.42) | $11.12 ($4.40–26.06) | $5.61 ($2.31–14.29) | $3.82 ($1.54–9.79) |
| Lao PDR | LMI | WPR | $28.80 ($8.37–88.91) | $15.71 ($5.07–45.94) | $25.43 ($8.39–68.46) | $18.04 ($5.91–48.28) | $6.96 ($2.72–18.78) | $4.78 ($1.82–12.71) |
| Lebanon | LMI | EMR | $20.43 ($4.66–79.34) | $11.14 ($2.71–42.03) | $22.56 ($7.39–57.80) | $16.04 ($5.28–40.86) | $4.82 ($1.35–20.31) | $3.30 ($0.94–13.33) |
| Lesotho | LMI | AFR | $45.20 ($8.00–222.44) | $24.87 ($4.75–116.01) | $31.72 ($8.63–102.09) | $22.48 ($6.16–71.57) | $1.52 ($0.30–10.45) | $1.04 ($0.20–6.83) |
| Liberia | LI | AFR | $6.86 ($2.46–18.45) | $3.74 ($1.47–9.31) | $19.42 ($7.89–43.70) | $13.79 ($5.70–31.05) | $6.17 ($2.62–15.06) | $4.24 ($1.75–10.42) |
| Libya | UMI | EMR | $4.62 ($1.77–11.78) | $2.52 ($1.05–6.10) | $20.67 ($7.82–49.00) | $14.66 ($5.63–34.63) | $5.84 ($2.43–14.71) | $4.02 ($1.58–10.06) |
| North Macedonia | UMI | EUR | $28.10 ($9.82–77.58) | $15.37 ($5.89–39.05) | $22.51 ($8.50–53.97) | $15.96 ($6.06–38.02) | $7.42 ($2.89–19.81) | $5.08 ($1.95–13.44) |
| Madagascar | LI | AFR | $4.34 ($1.14–14.93) | $2.38 ($0.64–8.09) | $24.53 ($8.42–62.59) | $17.48 ($6.11–44.09) | $3.30 ($0.98–14.36) | $2.25 ($0.68–9.18) |
| Malawi | LI | AFR | $5.70 ($2.46–13.90) | $3.13 ($1.44–7.11) | $17.78 ($7.59–38.46) | $12.63 ($5.43–27.20) | $3.33 ($1.50–7.88) | $2.26 ($1.02–5.48) |
| Malaysia | UMI | WPR | $29.95 ($12.88–71.38) | $16.38 ($7.85–35.76) | $18.84 ($7.26–44.13) | $13.40 ($5.17–31.48) | $6.50 ($2.80–15.81) | $4.45 ($1.92–10.76) |
| Maldives | UMI | SEAR | $35.15 ($12.01–101.72) | $19.20 ($7.42–49.87) | $15.66 ($6.24–36.42) | $11.12 ($4.40–26.06) | $0.80 ($0.13–5.64) | $0.55 ($0.09–3.94) |
| Mali | LI | AFR | $3.41 ($1.53–8.08) | $1.87 ($0.87–4.26) | $20.89 ($7.69–50.88) | $14.83 ($5.55–36.13) | $4.42 ($2.07–10.58) | $3.02 ($1.43–7.30) |
| Marshall Islands | UMI | WPR | $35.66 ($9.35–124.27) | $19.47 ($5.73–63.81) | $23.83 ($8.58–59.41) | $16.96 ($6.07–41.55) | $4.75 ($1.83–13.15) | $3.27 ($1.25–9.03) |
| Mauritania | LMI | AFR | $16.76 ($6.33–43.06) | $9.14 ($3.77–21.76) | $21.45 ($8.38–49.62) | $15.22 ($6.03–34.80) | $7.37 ($2.94–19.23) | $5.04 ($1.97–13.08) |
| Mauritius | UMI | AFR | $38.63 ($12.22–116.80) | $20.95 ($7.55–58.44) | $18.84 ($7.26–44.13) | $13.40 ($5.17–31.48) | $2.43 ($0.74–8.47) | $1.65 ($0.49–5.96) |
| Mexico | UMI | AMR | $35.93 ($8.87–128.98) | $19.66 ($5.17–68.50) | $31.72 ($8.63–102.09) | $22.48 ($6.16–71.57) | $4.20 ($1.59–12.70) | $2.87 ($1.18–8.13) |
| Micronesia | LMI | WPR | $46.26 ($7.56–245.77) | $25.63 ($4.51–125.91) | $31.12 ($8.66–97.27) | $22.08 ($6.22–68.39) | $4.51 ($1.54–15.05) | $3.09 ($1.06–9.94) |
| Moldova | UMI | EUR | $39.57 ($10.83–128.38) | $21.53 ($6.50–65.40) | $24.50 ($8.32–64.45) | $17.40 ($5.91–45.42) | $6.39 ($2.52–17.00) | $4.38 ($1.72–11.56) |
| Mongolia | UMI | WPR | $32.34 ($12.73–82.82) | $17.60 ($7.84–40.29) | $16.93 ($6.68–38.86) | $11.95 ($4.73–27.86) | $8.91 ($3.25–23.74) | $6.09 ($2.20–16.35) |
| Montenegro | UMI | EUR | $48.03 ($9.42–216.92) | $26.46 ($5.65–111.82) | $29.45 ($8.50–88.77) | $20.90 ($6.15–63.00) | $6.29 ($2.38–18.22) | $4.32 ($1.60–12.03) |
| Morocco | LMI | EMR | $27.68 ($12.69–63.12) | $15.15 ($7.75–31.99) | $16.93 ($6.68–38.86) | $11.95 ($4.73–27.86) | $6.75 ($2.93–16.30) | $4.62 ($1.99–10.87) |
| Mozambique | LI | AFR | $4.80 ($1.25–16.53) | $2.63 ($0.70–8.96) | $24.86 ($8.47–63.51) | $17.68 ($6.18–44.55) | $3.82 ($1.39–12.73) | $2.60 ($0.97–8.19) |
| Myanmar | LMI | SEAR | $13.22 ($3.11–49.89) | $7.22 ($1.77–26.74) | $27.74 ($8.86–75.27) | $19.66 ($6.42–53.34) | $3.93 ($1.33–13.78) | $2.68 ($0.94–8.96) |
| Namibia | LMI | AFR | $29.21 ($6.54–116.95) | $16.06 ($3.86–61.75) | $27.37 ($8.85–73.84) | $19.45 ($6.42–52.26) | $5.75 ($1.96–18.81) | $3.92 ($1.34–12.60) |
| Nepal | LMI | SEAR | $11.92 ($5.83–26.35) | $6.56 ($3.57–13.14) | $15.46 ($6.41–34.78) | $11.01 ($4.58–24.57) | $5.32 ($2.38–12.85) | $3.64 ($1.60–8.67) |
| Nicaragua | LMI | AMR | $30.70 ($10.67–83.90) | $16.77 ($6.49–42.59) | $21.84 ($7.95–53.87) | $15.60 ($5.66–38.02) | $7.28 ($2.86–18.90) | $4.97 ($1.92–12.73) |
| Niger | LI | AFR | $1.77 ($0.72–4.57) | $0.97 ($0.41–2.37) | $18.06 ($7.12–42.81) | $12.81 ($5.15–30.16) | $6.88 ($2.56–20.80) | $4.68 ($1.78–14.05) |
| Nigeria | LMI | AFR | $2.10 ($0.87–5.29) | $1.15 ($0.48–2.92) | $23.31 ($8.06–59.66) | $16.60 ($5.85–42.18) | $1.63 ($0.37–9.48) | $1.12 ($0.27–6.09) |
| Pakistan | LMI | EMR | $40.22 ($11.83–129.07) | $21.97 ($7.16–65.52) | $21.84 ($7.95–53.87) | $15.60 ($5.66–38.02) | $7.03 ($2.77–18.22) | $4.80 ($1.87–12.29) |
| Papua New Guinea | LMI | WPR | $6.57 ($3.49–13.65) | $3.61 ($1.97–7.20) | $18.78 ($7.71–41.89) | $13.31 ($5.60–29.71) | $2.41 ($0.99–6.99) | $1.65 ($0.72–4.42) |
| Paraguay | UMI | AMR | $25.22 ($6.18–92.34) | $13.82 ($3.53–49.40) | $27.45 ($8.84–73.90) | $19.49 ($6.43–52.53) | $4.52 ($1.15–22.06) | $3.09 ($0.79–14.32) |
| Peru | UMI | AMR | $41.28 ($9.55–156.24) | $22.50 ($5.70–81.66) | $30.55 ($8.52–94.82) | $21.64 ($6.16–67.07) | $6.55 ($2.42–19.39) | $4.50 ($1.63–12.81) |
| Philippines | LMI | WPR | $37.83 ($9.67–131.60) | $20.72 ($5.66–69.19) | $31.63 ($8.45–106.72) | $22.60 ($6.14–75.73) | $5.74 ($2.30–15.78) | $3.94 ($1.61–10.51) |
| Rwanda | LI | AFR | $31.18 ($7.36–115.79) | $17.03 ($4.23–62.04) | $30.14 ($8.74–89.34) | $21.30 ($6.33–63.38) | $1.94 ($0.60–8.25) | $1.32 ($0.43–5.17) |
| Samoa | UMI | WPR | $6.56 ($2.73–16.02) | $3.60 ($1.59–7.98) | $14.49 ($5.98–34.27) | $10.33 ($4.29–24.39) | $2.75 ($0.96–8.38) | $1.88 ($0.64–5.75) |
| Sao Tome and Principe | LMI | AFR | $44.66 ($10.51–170.46) | $24.46 ($6.43–87.52) | $25.43 ($8.39–68.46) | $18.04 ($5.91–48.28) | $6.39 ($2.51–17.32) | $4.38 ($1.70–11.70) |
| Senegal | LMI | AFR | $41.29 ($10.64–147.41) | $22.64 ($6.50–75.58) | $23.59 ($8.25–60.64) | $16.75 ($5.89–42.41) | $4.81 ($1.89–13.12) | $3.31 ($1.28–8.98) |
| Serbia | UMI | EUR | $12.22 ($5.90–26.07) | $6.69 ($3.57–13.14) | $16.14 ($8.49–31.42) | $11.44 ($6.01–22.57) | $9.00 ($3.42–26.36) | $6.12 ($2.37–17.52) |
| Sierra Leone | LI | AFR | $33.25 ($12.72–85.32) | $18.12 ($7.87–42.10) | $17.52 ($6.84–40.32) | $12.41 ($4.87–28.93) | $7.81 ($2.95–20.67) | $5.33 ($1.97–14.34) |
| Solomon Islands | LMI | WPR | $2.59 ($1.01–6.85) | $1.42 ($0.59–3.55) | $15.00 ($6.10–35.79) | $10.69 ($4.43–25.20) | $5.76 ($1.92–17.21) | $3.97 ($1.24–11.92) |
| Somalia | LI | EMR | $27.21 ($7.59–91.58) | $14.87 ($4.52–46.56) | $23.59 ($8.25–60.64) | $16.75 ($5.89–42.41) | $7.39 ($2.88–19.47) | $5.05 ($1.95–13.17) |
| South Africa | UMI | AFR | $2.11 ($0.77–5.85) | $1.15 ($0.43–3.14) | $23.31 ($8.06–59.66) | $16.60 ($5.85–42.18) | $3.80 ($1.22–14.26) | $2.60 ($0.83–9.27) |
| South Sudan | LI | AFR | $27.53 ($6.99–98.23) | $15.10 ($3.95–51.96) | $28.47 ($8.82–78.71) | $20.10 ($6.41–55.92) | $4.31 ($1.53–14.18) | $2.94 ($1.10–9.21) |
| Sri Lanka | LMI | SEAR | $3.63 ($1.31–10.09) | $1.98 ($0.74–5.40) | $23.31 ($8.06–59.66) | $16.60 ($5.85–42.18) | $4.72 ($1.61–15.63) | $3.25 ($1.06–10.66) |
| St. Lucia | UMI | AMR | $28.50 ($12.66–66.10) | $15.50 ($7.76–33.46) | $16.29 ($6.47–37.71) | $11.53 ($4.57–26.84) | $4.25 ($1.66–11.31) | $2.90 ($1.10–7.89) |
| St. Vincent and the Grenadines | UMI | AMR | $35.15 ($12.01–101.72) | $19.20 ($7.42–49.87) | $15.66 ($6.24–36.42) | $11.12 ($4.40–26.06) | $5.04 ($1.91–13.83) | $3.44 ($1.28–9.51) |
| Sudan | LI | EMR | $35.15 ($12.01–101.72) | $19.20 ($7.42–49.87) | $15.66 ($6.24–36.42) | $11.12 ($4.40–26.06) | $5.37 ($2.06–14.54) | $3.66 ($1.37–10.04) |
| Suriname | UMI | AMR | $8.09 ($2.10–27.66) | $4.42 ($1.18–15.04) | $26.13 ($8.74–68.01) | $18.57 ($6.24–47.79) | $3.37 ($0.92–15.74) | $2.29 ($0.66–10.06) |
| Eswatini | LMI | AFR | $53.70 ($7.57–314.69) | $29.67 ($4.59–164.16) | $31.72 ($8.63–102.09) | $22.48 ($6.16–71.57) | $5.26 ($1.50–21.37) | $3.60 ($1.05–14.02) |
| Syrian Arab Republic | LI | EMR | $14.06 ($2.15–78.16) | $7.72 ($1.20–41.82) | $31.72 ($8.63–102.09) | $22.48 ($6.16–71.57) | $4.23 ($1.42–14.41) | $2.89 ($1.00–9.56) |
| Tajikistan | LMI | EUR | $11.72 ($5.42–26.94) | $6.43 ($3.31–13.31) | $15.33 ($6.39–34.48) | $10.94 ($4.57–24.35) | $7.82 ($2.98–20.57) | $5.34 ($2.01–14.25) |
| Tanzania | LMI | AFR | $9.66 ($3.60–24.88) | $5.28 ($2.06–13.21) | $24.84 ($7.84–70.42) | $17.66 ($5.67–49.47) | $4.50 ($1.95–11.77) | $3.08 ($1.43–7.49) |
| Thailand | UMI | SEAR | $29.90 ($12.84–70.60) | $16.33 ($7.73–35.82) | $19.49 ($7.54–45.82) | $13.91 ($5.28–32.97) | $5.66 ($2.58–13.39) | $3.88 ($1.80–8.99) |
| Timor-Leste | LMI | SEAR | $11.33 ($3.45–35.41) | $6.19 ($2.02–17.91) | $22.46 ($8.49–53.56) | $15.95 ($6.14–37.03) | $5.74 ($2.39–14.52) | $3.93 ($1.61–10.07) |
| Togo | LI | AFR | $6.06 ($2.77–13.27) | $3.29 ($1.68–6.80) | $16.78 ($7.22–36.84) | $11.95 ($5.18–26.02) | $5.49 ($2.30–13.53) | $3.74 ($1.53–9.39) |
| Tonga | UMI | WPR | $35.15 ($12.01–101.72) | $19.20 ($7.42–49.87) | $15.66 ($6.24–36.42) | $11.12 ($4.40–26.06) | $6.81 ($2.62–17.86) | $4.65 ($1.75–12.47) |
| Tunisia | LMI | EMR | $29.72 ($12.72–71.44) | $16.19 ($7.82–35.58) | $16.29 ($6.47–37.71) | $11.53 ($4.57–26.84) | $7.83 ($2.96–20.73) | $5.35 ($1.98–14.39) |
| Turkiye | UMI | EUR | $25.55 ($12.09–59.12) | $14.00 ($7.24–30.23) | $15.66 ($6.24–36.42) | $11.12 ($4.40–26.06) | $5.89 ($2.67–14.07) | $4.02 ($1.85–9.16) |
| Turkmenistan | UMI | EUR | $18.58 ($7.40–47.84) | $10.16 ($4.48–24.20) | $13.56 ($5.76–30.69) | $9.69 ($4.13–21.83) | $8.78 ($3.32–22.93) | $6.01 ($2.22–15.80) |
| Tuvalu | UMI | WPR | $35.49 ($11.53–104.30) | $19.36 ($7.15–52.00) | $18.19 ($7.06–42.05) | $12.91 ($5.04–30.17) | $4.56 ($1.72–12.56) | $3.11 ($1.14–8.70) |
| Uganda | LI | AFR | $7.82 ($3.83–16.90) | $4.31 ($2.26–8.79) | $19.36 ($6.95–55.85) | $13.82 ($5.04–38.24) | $2.65 ($0.78–8.50) | $1.83 ($0.50–5.82) |
| Ukraine | UMI | EUR | $34.19 ($12.01–91.82) | $18.62 ($7.25–46.36) | $22.70 ($8.11–57.12) | $16.16 ($5.79–39.99) | $6.87 ($2.72–17.85) | $4.69 ($1.86–12.02) |
| Uzbekistan | LMI | EUR | $28.52 ($11.83–69.22) | $15.60 ($7.05–34.97) | $21.05 ($7.81–51.13) | $14.99 ($5.52–36.33) | $5.90 ($2.69–13.90) | $4.03 ($1.90–9.20) |
| Vanuatu | LMI | WPR | $53.51 ($7.56–313.17) | $29.56 ($4.58–162.84) | $31.72 ($8.63–102.09) | $22.48 ($6.16–71.57) | $5.36 ($1.63–20.36) | $3.66 ($1.13–13.53) |
| Venezuela | UMI | AMR | $36.36 ($7.63–149.06) | $19.86 ($4.44–79.90) | $30.77 ($8.70–94.29) | $21.82 ($6.27–66.53) | $4.18 ($1.05–20.49) | $2.85 ($0.73–13.54) |
| Viet Nam | LMI | WPR | $23.92 ($11.41–55.28) | $13.13 ($6.81–28.14) | $16.29 ($6.47–37.71) | $11.53 ($4.57–26.84) | $3.70 ($1.75–8.72) | $2.55 ($1.23–5.74) |
| Yemen | LI | EMR | $7.09 ($1.33–32.16) | $3.87 ($0.74–17.44) | $27.64 ($8.86–74.77) | $19.60 ($6.43–53.14) | $3.24 ($0.85–16.15) | $2.21 ($0.61–10.20) |
| Zambia | LMI | AFR | $8.30 ($3.60–19.77) | $4.55 ($2.10–10.09) | $17.18 ($7.42–37.00) | $12.21 ($5.32–25.99) | $6.82 ($3.14–15.69) | $4.66 ($2.14–10.42) |
| Zimbabwe | LMI | AFR | $14.93 ($6.96–32.79) | $8.17 ($4.22–16.48) | $17.21 ($7.43–37.04) | $12.23 ($5.32–26.12) | $8.48 ($3.98–19.19) | $5.76 ($2.74–13.33) |

^a^ LI: Gross national income (GNI) per capita of $1,135 or less in 2024; LMI: GNI per capita $1,136 and $4,495; UMI: GNI per capita of $4,496 and $13,935 (World Bank 2024) [18].

Note: AFR = African region; AMR = Region of the Americas; DTP3 = diphtheria-tetanus-pertussis third dose coverage; EMR = Eastern Mediterranean region; EUR = European region; GDP = gross domestic product; LI = low-income; LMI = lower middle-income; SEAR = Southeast Asian region; UMI = upper middle-income; WPR = Western Pacific region.
